# HAT-field: a cheap, robust and quantitative point-of-care serological test for Covid-19

**DOI:** 10.1101/2022.01.14.22268980

**Authors:** Etienne Joly, Agnès Maurel Ribes

## Abstract

We have recently described a very simple and cheap serological test called HAT to detect antibodies directed against the RBD of the SARS-Cov-2 virus. HAT is based on hemagglutination, triggered by a single reagent (IH4-RBD) comprised of the viral RBD domain fused to a nanobody specific for glycophorin, which is expressed at very high levels at the surface of human red blood cells (RBCs).

One of the main initial goals of this study was to devise a test protocol that would be sensitive and reliable, yet require no specialized laboratory equipment such as adjustable pipets, so that it could be performed in the most remote corners of the world by people with minimal levels of training. Because antibody levels against the viral RBD have been found to correlate closely with sero-neutralisation titers, and thus with protection against reinfection, it has become obvious during the course of this study that making this test reliably quantitative would be a further significant advantage.

Using IH4-RBD based on the original Wuhan sequence, we have found that, in PBN, a buffer which contains BSA and sodium azide, the reagent is stable for over 6 months at room temperature, and that PBN also improves HAT performance compared to using straight PBS. We also show that performing HAT at either 4°C, room temperature or 37°C has minimal influence on the results, and that quantitative evaluation of the levels of antibodies directed against the SARS-CoV-2 RBD can be achieved in a single step using titration of the IH4-RBD reagent.

The HAT-field protocol described here requires only very simple disposable equipment and a few microliters of whole blood, such as can be obtained by finger prick. Because it is based on a single soluble reagent, the test can be adapted very simply and rapidly to detect antibodies against variants of the SARS-CoV-2, or conceivably against different pathogens. HAT-field appears well suited to provide quantitative assessments of the serological protection of populations as well as individuals, and given its very low cost, the stability of the IH4-RBD reagent in the adapted buffer, and the simplicity of the procedure, could be deployed pretty much anywhere, including in the poorest countries and the most remote corners of the globe.

**Note: This manuscript has been refereed** by Review Commons, and modified thanks to the comments and suggestions from three referees. Those comments, and our replies, are provided at the end of the manuscript’s pdf, and can also be accessed by clicking on the blue tab found to the right of the MedRXiv window.

## Introduction

For the past two years, the Covid-19 pandemic has preoccupied the whole world, and it remains a major concern for all nations, albeit with different perspectives depending on their wealth. In affluent nations, most people have now been vaccinated, and the main issues now are when to start offering booster vaccinations and to whom. Poorer countries, by contrast, have had limited access to vaccines or even to diagnostic tests simply to follow the progress of the pandemic within their populations. To date, most tests available to monitor immune responses against SARS-CoV-2 either require elaborate laboratory procedures and equipment, or are not sufficiently sensitive or quantitative to be of real value (Abbasi, 2021; Moshe et al., 2021; Ong et al., 2021). For both affluent and less affluent countries, access to a robust and reliably quantitative point-of-care (PoC) serological test would be a great asset to tackle these problems. Such a test would allow health professionals, and health authorities, to distinguish people with either no or waning levels of antibodies, who should have priority for vaccination or re-vaccination, from those with high levels of antibodies against the SARS-CoV-2 virus, who may not need to be vaccinated or revaccinated immediately, and may actually be the ones most likely to suffer undesirable effects from vaccine injections.

Last year, we described a very simple, inexpensive serological test for Covid-19 called the HAT (hemagglutination test; (Townsend et al., 2021). HAT uses a recombinant protein (IH4-RBD) comprised of a nanobody, IH4, which binds to human glycophorin at the surface of red blood cells (Habib et al., 2013), fused to the receptor-binding domain (RBD) of the SARS-CoV-2 virus. When mixed with diluted human blood, this reagent coats the red blood cells (RBCs) and, if antibodies to the viral RBD domain are present in the blood sample, they will cause hemagglutination. This test thus detects specifically antibodies against the RBD, which means that it can be used as a surrogate sero-neutralization test since those antibodies are the main ones endowed with sero-neutralizing activity against the virus (Ertesvåg et al., 2021; Jeewandara et al., 2021a; Lamikanra et al., 2021). Another important feature of HAT is that, because it is based on a soluble reagent, it can be adapted very easily and rapidly to detect antibodies against different variant forms of the virus (Ertesvåg et al., 2021; Jeewandara et al., 2021a) or presumably to other pathogens if needed be, for example in the context of a newly arising pathogen.

In the format initially described for HAT, quantitative evaluation of the levels of antibodies was possible via serial dilutions of serum or plasma before mixing with washed autologous RBCs, or obtained from O-donors (Townsend et al., 2021). In its simple single-point format, HAT was recently used to measure seropositivity rates in Sri Lanka and compared well to a sensitive ELISA (Jeewandara et al., 2021b). Here, we describe an adapted protocol, called HAT-field, which is quantitative through titration of the IH4-RBD reagent and can be performed in a single simple step with no specialized equipment. The observation that the performances of the assay are minimally affected by temperatures and that, in the optimized HAT-field buffer, which contains BSA and azide, the reagent is stable for weeks with no refrigeration required could also greatly facilitate the use of HAT-field in remote locations.

## Results

### BSA prevents adsorption of IH4-RBD to the reaction wells

Following the method originally described by Wegmann and Smithies (Wegmann and Smithies, 1966), the original HAT protocol uses 96 conical-well plates (Townsend et al., 2021). When appropriately diluted blood is mixed with the IH4-RBD reagent in these conical wells, the RBCs sediment during the incubation of 60 minutes; hemagglutination due to specific antibodies against RBD in the blood is observed by the formation of persistent ‘buttons’ of RBCs in the bottom of the well when the plate is tilted, whereas in the absence of hemagglutination a ‘teardrop’ shape forms (Townsend et al., 2021).

To perform HAT in field conditions and/or on large numbers of samples, it would be much simpler for the users to be provided with the IH4-RBD reagent already distributed in the V-well plates used to perform the HAT tests. The plates, however, are made of polystyrene, to which many proteins tend to adsorb. (This non-specific adsorption is the basis for many ELISA tests; (Kenny and Dunsmoor, 1983). Indeed, when the IH4-RBD reagent was diluted in PBS and placed in the wells, some of it was readily adsorbing to the plastic of the wells’ slopes, and causing the formation of diffuse veils in hemagglutinated wells, which not only raised concerns about losing some of the active reagent by its immobilization on the plastic, but could make the reading of the results of the HAT tests less clear than the formation of bright red buttons (Figure 1A). Furthermore, in preliminary experiments involving serial dilutions of the IH4-RBD reagent, we found that the diluted IH4-RBD reagent tended to be lost rapidly through this phenomenon of adsorption.

**Figure 1:**
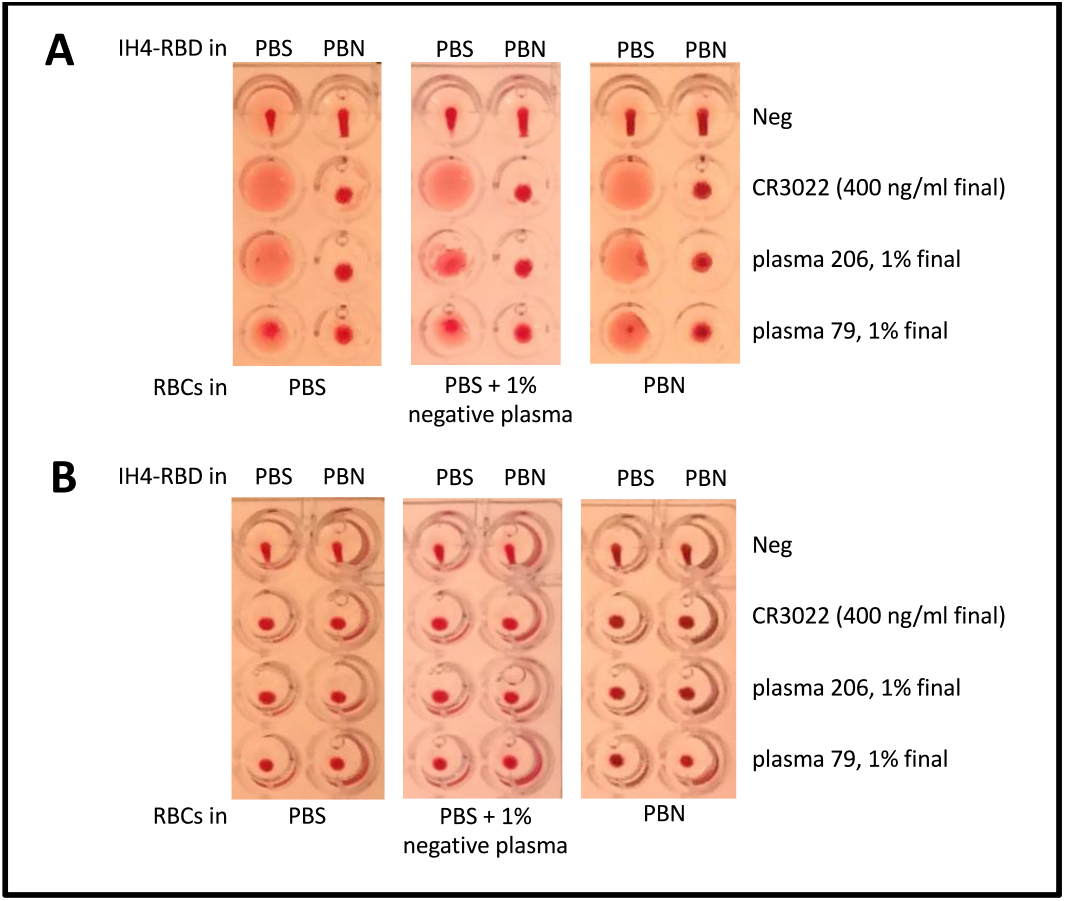
BSA prevents adsorption of IH4-RBD to the reaction wells. A) HAT was performed in uncoated wells prefilled with IH4-RBD reagent diluted in either PBS (left columns) or PBS + 1% BSA + 3mM sodium azide (PBN, right columns). RBCs, resuspended in either PBS (left panels), in PBS supplemented with 1% seronegative autologous plasma (middle panels), or PBN (right panels) and various antibodies against SARS-CoV-2 were added to each well to test for hemagglutination. In the absence of antibody (Neg) the typical teardrop structure can be seen in each well. In the presence of a monoclonal antibody against RBD (CR3022), or plasma from convalescent Covid-19 patients, a veil structure forms in the absence of BSA, whereas a button forms when BSA is present. (see Methods for details) B) HAT as in (A) but performed in wells precoated with BSA. In the presence of the monoclonal antibody or convalescent patient plasma, hemagglutination is observed as a button rather than the veils seen in (A). Similar data were obtained from three experiments.

We solved this problem of veil formation by diluting the IH4-RBD reagent in PBN (Figure 1A, right columns of each panel), which is PBS containing 1% BSA and 3mM sodium azide, to prevent contamination by micro-organisms. In PBN, rather than veils, buttons of hemagglutination were observed and could be distinguished easily from the teardrops in the negative controls. These buttons formed whether the RBCs, which were added after the IH4-RBD reagent, were resuspended in PBS, in PBS supplemented with 1% plasma from the same seronegative donor as the RBCs, or in PBN. We conclude from this experiment that veil formation is due to adsorption of the IH4-RBD reagent to the polystyrene walls of the wells. This interpretation is supported by our finding that no veils formed when the wells were precoated with BSA and rinsed with PBS before performing the assay (Figure 1B).

### Azide and BSA increase the sensitivity of HAT

To investigate whether the presence of BSA and azide diminishes the performance of the HAT, we diluted the IH4-RBD reagent in PBS, in PBS containing 3mM sodium azide (PBS-N3), or in PBN and used these diluted reagents to test hemagglutination of whole blood and O-RBCs from a seronegative donor resuspended in 1% plasma from the same donor, in the presence of serial dilutions of either the monoclonal anti-RBD CR3022 or of an immune serum (Figure 2).

**Figure 2:**
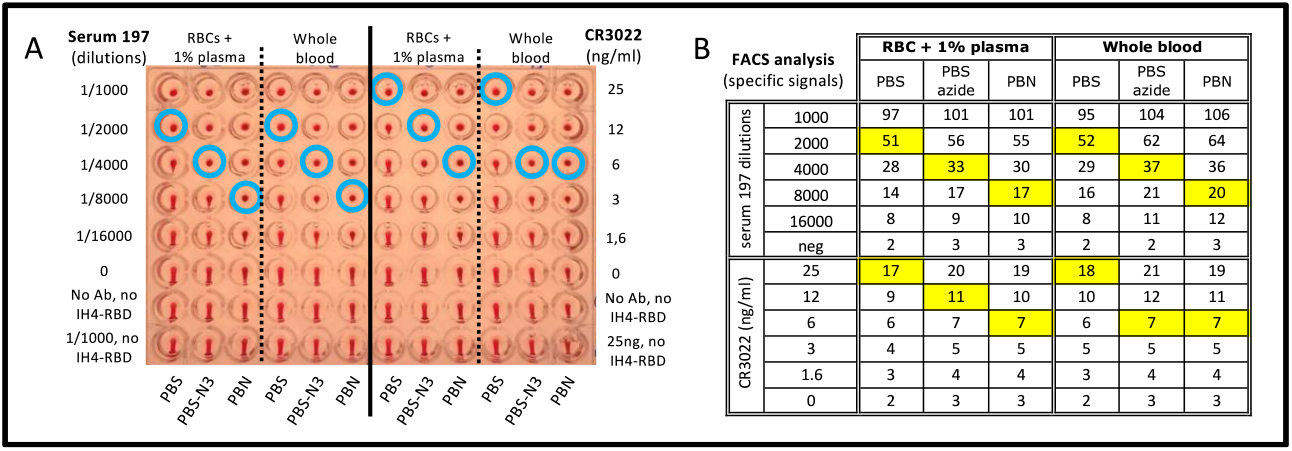
Azide and BSA increase the sensitivity of HAT. A) The effects of azide and BSA on HAT performed using IH4-RBD at 1 μg/ml final concentration, and RBCs from a seronegative donor resuspended in PBS, PBS-N3 or PBN and 1% plasma from the same donor, or with whole blood from the same donor, and with serial dilutions of an immune serum from a convalescent Covid-19 patient (Serum 197; left columns) or a monoclonal anti-RBD (CR3022; right columns). The three negative controls were: no antibody (0), neither antibody nor IH4-RBD, and the most concentrated serum or antibody condition with no IH4-RBD. Blue circles indicate the titration endpoints. (see Methods for details) B) After HAT, the RBCs were resuspended, stained with a fluorescent secondary anti-human Ig antibody and analyzed by FACS. The numbers shown correspond to specific signals, i.e. the difference in GMFI values of each sample with that of the control sample incubated in the same buffer with no antibody or IH4-RBD. The squares highlighted in yellow indicate the titration endpoints. Comparable results were obtained in four similar experiments.

Rather than diminishing the performance of HAT, we found that the presence of azide improved its sensitivity: when HAT was performed in PBS-N3 rather than PBS, the titration endpoints (figure 2A, blue circles) were shifted by one double dilution (DD), and this occurred both with immune sera and with monoclonal antibodies. Addition of 1% BSA sometimes improved sensitivity by another DD, but we only saw this in some experiments, and not others.

To investigate the possible cause of the increased sensitivity of HAT in the presence of sodium azide, we used flow cytometry (FACS) to analyze the amount of antibody bound to the RBCs at the end of the HAT. Dilution of the IH4-RBD reagent in PBS-N3 or in PBN resulted in a small increase in the amount of antibody bound to the surface of the RBCs, but not to an extent that would explain the increased sensitivity (Figure 2B). We postulate that the increased sensitivity due to the presence of azide may, instead, be primarily due to an ‘ageing’ effect on the RBCs.

### IH4-RBD is very stable when diluted in PBN

For use in the field, it would be most convenient for the IH4-RBD reagent to be pre-distributed in the wells, raising the question of the stability of working dilutions of the reagent, both in the cold and at ambient temperatures. To investigate the stability of IH4-RBD, we prepared aliquots of IH4-RBD at 2 μg/ml in either PBS-N3 or in PBN, on various dates over the course of 15 months and stored those aliquots at 4°C, room temperature (RT) or 37°C. Those IH4-RBD aliquots of various ages were then used to perform HAT titrations of the reagent in the presence of constant amounts of the CR3022 monoclonal antibody (Figure 3). In some experiments, to evaluate more precisely the remaining activity of IH4-RBD after incubation, we also quantified the amounts of antibodies bound to the surface of the RBCs by using FACS (red numbers in Figure 3).

**Figure 3:**
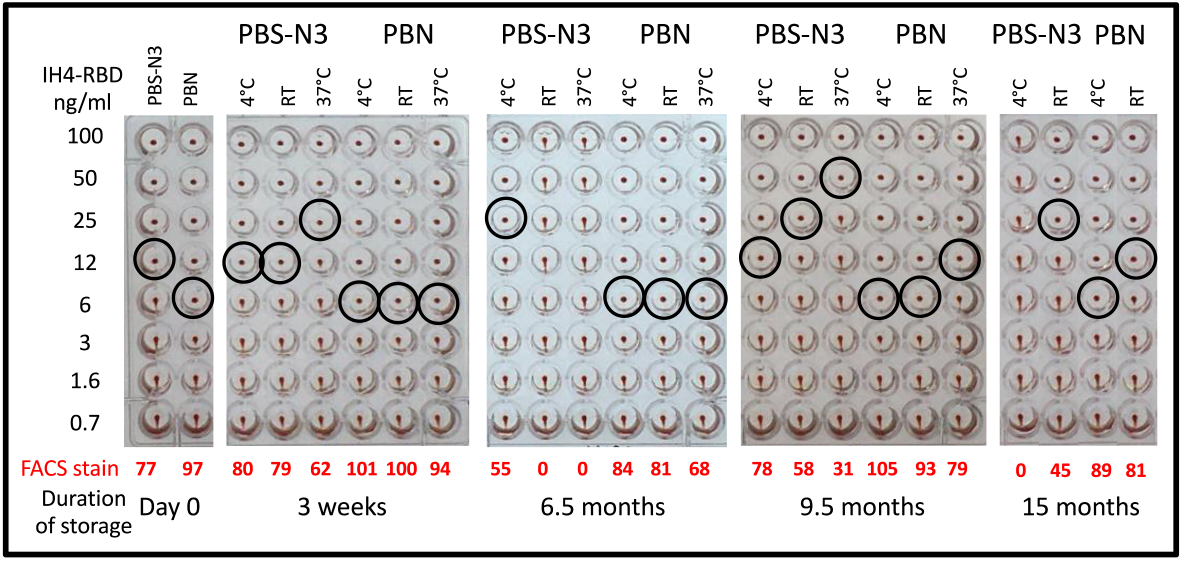
IH4-RBD is very stable when diluted in PBN. The effects of long-term storage of IH4-RBD in PBS-N3 or PBN at 4°C, RT and 37°C, as determined by HAT and by FACS analysis of antibody binding to RBCs after HAT. Hemagglutination end-points in the presence of the CR3022 monoclonal antibody at 100 ng/ml (black circles) were determined by titration of aliquots of working dilutions of IH4-RBD (2 μg/ml) stored for up to 15 months at the indicated temperatures. The red numbers indicate the intensity of the specific fluorescent staining recorded by FACS analysis performed after the HAT assay on the RBCs from the samples incubated with 100 ng/ml IH4-RBD (see Methods for details). Similar data were obtained in seven experiments, some of which also included using diluted sera from convalescent patients in parallel to the CR3022 monoclonal antibody (not shown).

By performing such experiments repeatedly, we found that IH4-RBD is remarkably stable when diluted in PBN: no significant loss of activity was seen for the IH4-RBD PBN dilution kept for 15 months at 4°C, and this was true for up to 6.5 months at room temperature. At 37°C, we did see some progressive loss of activity, but this only resulted in the loss of one DD in HAT sensitivity at 9.5 months (after 15 months, evaporation had caused the loss of what was left of the aliquots kept at 37°C).

On the other hand, the activity of IH4-RBD dilutions prepared in PBS-azide were usually already lower by one DD than those prepared in PBN on day zero. Furthermore, we observed marked variability over time between the IH4-RBD dilutions prepared in PBS-azide on different dates: some batches showed a drop of just one DD compared to the dilutions prepared in PBN, and stayed stable for many weeks after this; for others, however, we witnessed much more marked losses over time, dropping to undetectable levels after just a few weeks, even for tubes kept at 4°C. In retrospect, we suspect that this variability may be linked to the fact that, because the Covid-19 crisis had caused a penury of plasticware, different types and brands of plastic tubes had to be used to prepare and stock the IH4-RBD dilutions on different dates, and those different tubes probably had different protein-binding capacities, resulting in the variable loss of the diluted IH4-RBD protein.

The important take-home message we draw from this set of experiments is that, regardless of the brand or type of plastic tubes used, as long as IH4-RBD was diluted in PBN, the activity of the diluted stocks was always remarkably reproducible, and stable for over a year if kept at 4°C, and with only marginal losses for dilutions kept at room temperature or 37°C. This remarkable stability of IH4-RBD, which is the sole reagent required for HAT, could greatly facilitate making this serological test available to populations living in remote environments, with no access to refrigeration.

### Temperature has little influence on HAT results

For use as a PoC test in the field, the HAT should be robust in many environments and, in particular, at a broad range of temperatures. To evaluate how temperature influences the results of the HAT, we set-up three identical 96-wells plates for 2D titration experiments (i.e. double dilutions of antibodies in one direction, and of the IH4-RBD reagent in the other) and incubated those at three different temperatures: at 4°C (on ice in a cold room), at RT (*ca*. 21°C), and at 37°C (in a CO_2_ cell culture incubator; Figure 4).

**Figure 4:**
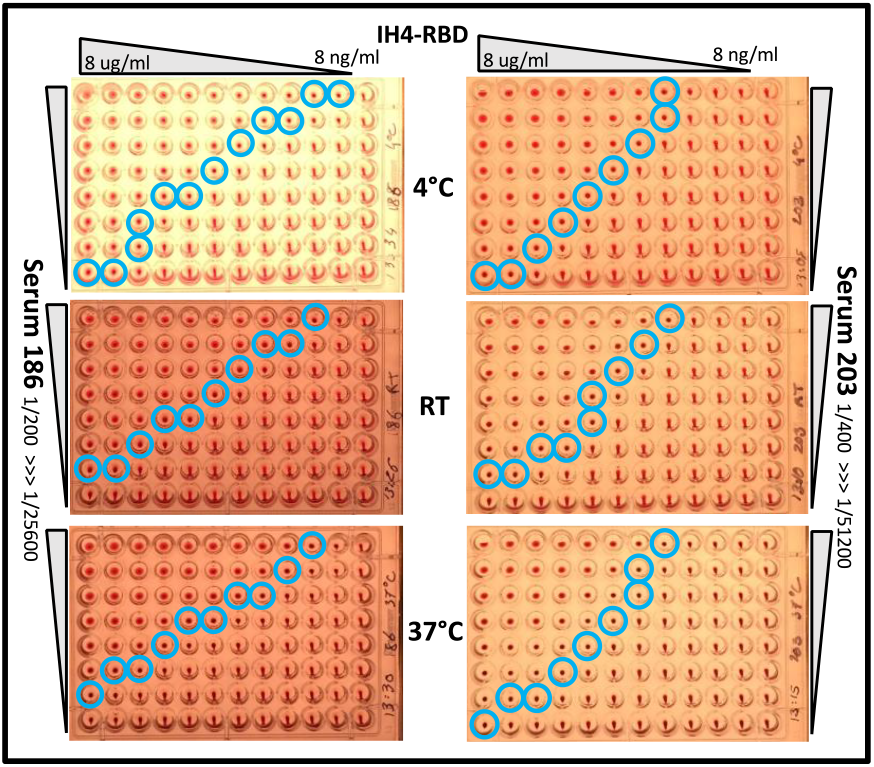
Temperature has little influence on HAT results. To determine the effect of temperature on the performance of HAT, three parallel plates were setup for 2D titrations, with DD of IH4-RBD going from 8 μg/ml to 8 ng/ml along lines, and DD of two different immune sera from convalescent Covid-19 patients down columns (sera 186 and 203, see Methods for practical details). After incubation at the indicated three temperatures, no substantial differences were seen in titration end-points (blue circles). Similar results were obtained in 3 independent experiments, using a total of 3 different immune sera, 2 plasmas, and the CR3022 monoclonal antibody.

Incubation temperature had little or no discernable influence on the hemagglutination endpoints (blue circles), with the possible exception of the wells containing the highest concentration of IH4-RBD and very diluted sera, where incubation at 4°C resulted in a small improvement in sensitivity when compared to the assays performed at RT or 37°C.

### The HAT-field protocol

The observation that, in 2D titrations such as those shown in Figure 4, the titration endpoints were distributed in an almost linear fashion on an X–Y axis suggested to us that a quantitative version of HAT might be developed by using dilutions of IH4-RBD rather than by using serial dilutions of plasma or sera and donor RBCs. We have now devised such a quantitative protocol, which only requires, for each test, one lancet, one plastic Pasteur pipet, one plastic tube containing 300 μl of PBS–2mM EDTA, 10 μl of whole blood, and one column of 8 conical wells on a 96-well plate, preloaded with 60 μl/well of a range of concentrations of IH4-RBD (Figure 5).

**Figure 5:**
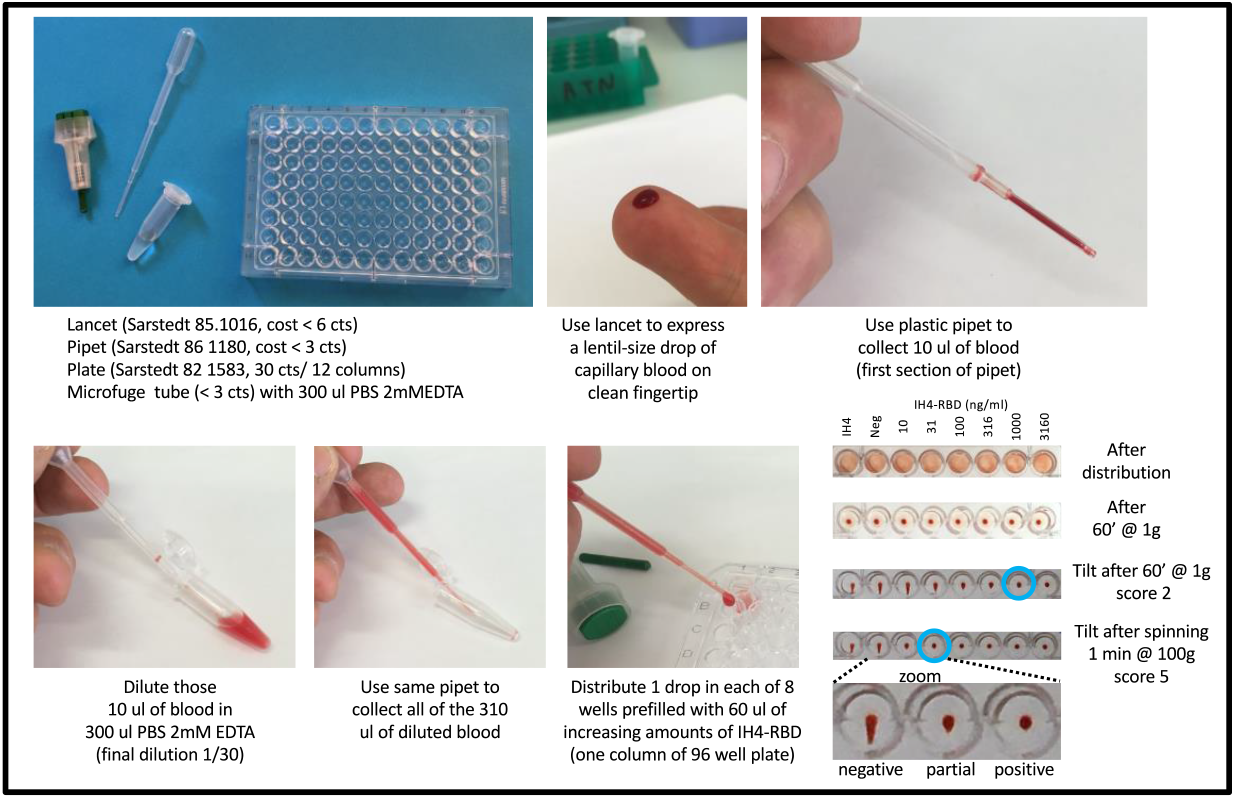
Schematic description of the HAT-field protocol.

The lancet is used to express a lentil-size drop of capillary blood from a clean fingertip of the subject to be tested. The plastic Pasteur pipet is used to collect 10 μl of that blood, which corresponds to filling the first section of the pipet (the precise volume of blood collected is not critical; it may vary by as much as 30% with no detectable influence on the results). The blood is diluted *ca*. thirty-fold in the tube containing 300 μl of PBS -2mM EDTA. The same pipet is then used to collect all 310 μl of this diluted blood and to transfer one drop into each of the 8 wells of a 96-well plate, prefilled with 60 μl of PBN containing 7 concentrations of IH4-RBD, and a negative control well containing either PBN or IH4 alone (not fused to RBD) diluted in PBN to a similar molar concentration as the highest IH4-RBD concentration used. As for the original HAT, the plate is incubated at room temperature, tilted after 60 minutes, and photographed after *ca*. 20 seconds. The photograph will later be used to score the samples. Scoring simply corresponds to the number of fully hemagglutinated wells in a column, and thus goes from 0 (no hemagglutination in the well with the highest concentration of IH4-RBD, i.e. 3.16 μg/ml) to 7 (full hemagglutination in the well with the lowest concentration of IH4-RBD, i.e. 3.16 ng/ml). For the tilting and the photographing, we find it convenient to use a very simple home-made light box ([see tutorial](https://www.youtube.com/watch?v=e5zBYd19nIA&feature=youtu.be)) and a standard smart phone camera.

One limitation of HAT is that it is not as sensitive as ELISA, CLIA (chemiluminescence immunoassay) or FACS (Lamikanra et al., 2021; Maurel Ribes et al., 2021). As we have seen above (Figure 4), increased sensitivity can be attained by the use of more IH4-RBD reagent, but we found that another way to increase HAT sensitivity was to perform prolonged incubations. After 5 hours, for example, we saw a very significant improvement in sensitivity, with the titration endpoints increasing for most samples by 2 or 3 dilution points when compared to those after 60 minutes, with fewer and fewer samples that were detected positively by FACS remaining below the threshold value of 1 for HAT-field (Figure S1, first line).

Such long incubations are, however, not practical for a test intended for use in field settings. To overcome this problem, we found that centrifugation of the plates at 100g for 1 minute increased sensitivity to a level equivalent, or even slightly superior to that of incubating the plates for 5 hours. This centrifugation step, moreover, may be performed 15 minutes after distributing the diluted blood in the plate, with similar results to those obtained if the plates were centrifuged after 60 minutes incubation (Figure S1, second line). With access to the means to centrifuge the assay wells (which can be achieved in adapted salad-spinners, see Discussion), the HAT-field protocol can thus be completed in less than 30 minutes, which would be compatible with performing it in certain field settings, for example in the context of vaccination centers, to identify individuals with high levels of antibodies, who might not need to be vaccinated or re-vaccinated.

### Validation of the HAT-field protocol

To validate the performance of the HAT-field protocol, we used a panel of 60 EDTA whole-blood samples collected in early September from patients in the hematology department of Toulouse University hospital. The samples were picked randomly from clinical samples left over after the prescribed hematology analyses had been performed. At that time, over 85% of the adult population had been vaccinated in France, and we thus expected a large proportion of the samples to be seropositive against the S protein of SARS-CoV-2, albeit at various levels.

The results obtained by HAT-field on these 60 whole blood samples were compared with those obtained by testing the plasma from the same samples by using the original HAT protocol with donor RBCs (Townsend et al., 2021), and by using the FACS-based Jurkat-S&R-flow test (Maurel Ribes et al., 2021), which is very sensitive, quantitative, and allows isotyping of the antibodies reacting against the S protein (Figure 6).

**Figure 6:**
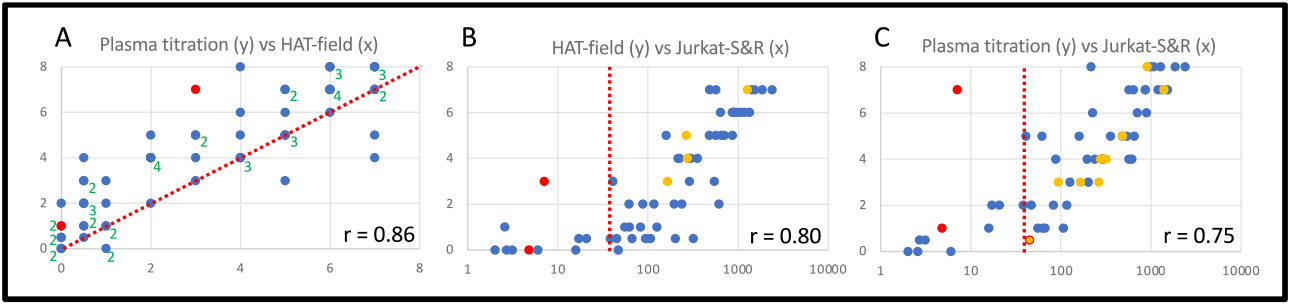
Validation of the HAT-field protocol by comparison with laboratory tests. Sixty randomly selected blood samples were used to compare the results of the HAT-field protocol with those of HAT plasma titrations and the Jurkat-S&R-flow test (see Methods). The graphs show one-on-one comparisons of the results obtained with those three tests, as indicated, using the values obtained by centrifuging the plates after 15 minutes (for HAT-field), and after 60 minutes for plasma titrations. Pearson’s correlation coefficients are indicated in the bottom right corners. On panel A, because of the discrete nature of the scales used for both the X and Y axes, many points overlap on top of one another. In such cases, their numbers are indicated by the adjacent green numbers. The dotted line in A indicates the position of the median, and those in B and C indicate the threshold for positive samples in the Jurkat-S&R-flow. The two red dots in each graph correspond to two negative samples, which gave false-positive results in HAT plasma titrations due to their reactivity against the IH4 nanobody moiety of the reagent. The orange dots correspond to samples positive in the Jurkat-S&R-flow test that showed some reactivity against the IH4 nanobody alone, albeit with lower titers than against the IH4-RBD reagent. For the sample represented by an orange dot surrounded by a red circle in C, the plasma titration against the nanobody alone was positive, but it led to only partial hemagglutination with the IH4-RBD reagent (for actual values, see sample 48 in data file).

The very good correlation between the results of all three tests validates that the HAT-field protocol can be used for quantitative assessment of the levels of antibodies contained in a whole-blood sample in a single step, without the sophisticated equipment needed for the Jurkat-S&R-flow test, and without needing to separate the RBCs from plasma or serum and having access to RBCs from an O-donor, as in the experiment using the original HAT to perform plasma titrations.

### HAT-field works with Delta-variant IH4-RBD

Our IH4-RBD reagent was designed to present the RBD sequence of the original Wuhan variant (residues 340–538 of the S protein; (Townsend et al., 2021). At the time of our study, however, a large proportion of the SARS-CoV-2 viruses circulating in France belonged to the Delta variant lineage ([see epidemiological report here](https://www.santepubliquefrance.fr/maladies-et-traumatismes/maladies-et-infections-respiratoires/infection-a-coronavirus/documents/bulletin-national/covid-19-point-epidemiologique-du-30-septembre-2021)), which has two mutations in the RBD domain (L452R and T478K in the B.1.617.2 strain) (Ertesvåg et al., 2021; Jayathilaka et al., 2021). We therefore wanted to compare the results obtained with the IH4-RBD-Wuhan reagent (used above) with a reagent that incorporates the two mutations in the Delta variant, IH4-RBD-Delta. We tested the 60 blood samples with the IH4-RBD-Wuhan and IH4-RBD-Delta reagents in both the HAT-field and original HAT plasma titration assays. For most samples, the scores obtained were one or two units higher when the Wuhan IH4-RBD reagent was used than those with the IH4-RBD-Delta reagent (Figure S3). This is consistent with a previous report that, in vaccinated people, HAT titers obtained with the IH4-RBD-Delta tend to be lower than with the IH4-RBD-Wuhan (Jayathilaka et al., 2021), and with the fact that, at the time of our study, most people in the French population had antibodies due to being vaccinated and not as a consequence of a previous infection by the SARS-CoV2 virus (retrospective analysis of clinical information on our cohort of 60 blood samples revealed that only three samples were from patients who had ever had a positive PCR test for SARS-CoV-2 (see data file and Figure S3).

## Discussion

In this paper, we describe an adaptation of the HAT protocol which is quantitative, shows satisfactory sensitivity, and can be used in the field with no specialized laboratory equipment such as adjustable pipets and disposable tips. This was made possible by the observations that i) the use of PBN results in markedly improved HAT robustness and sensitivity ii) HAT sensitivity is markedly improved by prolonged incubations (or by brief low-speed centrifugation), albeit with a parallel drop in specificity iii) quantification could be achieved by titrating the IH4-RBD reagent rather than the plasmas or sera. Incidentally, we realized recently that such an approach of titrating the RBC-binding reagent had been suggested previously for an HIV serodiagnostic test (Kemp et al., 1988).

PBN is a buffer containing both 1% BSA and azide, which has several concomitant advantages: i) Used as a dilution buffer for the IH4-RBD, it blocks the reagent’s nonspecific adsorption to plastic, and results in its much improved stability over time, even if kept out of the cold. ii) The conjoint action of azide and BSA results in slightly increased sensitivity, probably because they both improve the settling of the RBCs at the bottom of the wells. iii) In experiments which involve the use of O-RBCs, for example when using HAT to titrate plasmas or sera, the use of whole blood (or the addition of 1% seronegative plasma or serum) is no longer necessary when PBN is used to prepare a suspension of washed O-RBCs. We feel that this advantage is quite significant since most blood donors in the population have now become seropositive because of vaccinations.

Once we had found that the use of centrifugation, combined to that of PBN as a buffer, could result in a marked improvement of the sensitivity of the ‘standard’ HAT assay in test samples, we needed to validate the performance of the modified HAT assay on clinical samples. For this, we made use of a cohort of 60 clinical blood samples of unknown serological status, and characterized those using the Jurkat-S&R-flow test, which is both extremely sensitive, quantitative, and allows the semi-quantitative isotyping of the plasmatic antibodies (Maurel Ribes et al., 2021). As seen on figure 6, we found very good correlations between all three tests: HAT-field, HAT plasma titrations, and the Jurkat-S&R-flow test.

Whilst HAT was, from its initial conception, always intended primarily to be carried out on capillary blood obtained by fingertip pricks, because of regulatory restrictions, all the experiments described in this paper had to be performed on samples of venous blood collected by phlebotomy. A recent study has shown that, as could be expected, HAT performed on capillary blood gives the same results as on venous blood samples (Ertesvåg et al., 2021), and preliminary results which we have obtained recently suggest that the results of the HAT-field protocol performed on capillary blood indeed correlate just as well with those of the Jurkat-S&R-flow test as those obtained with venous blood (Joly et al., man. in prep.)

On the graph on the left of figure 6, which compares the scores of titrations by standard HAT to those obtained with HAT-field, most points are sitting above the median line, with 11 points not scoring positive by HAT-field (i.e. a score < 1) whilst being positive by standard HAT titration (i.e. score ≥ 1). In the conditions used in this study, the HAT-field protocol was thus markedly less sensitive than the optimized standard HAT approach, which can be explained by the conjunction of 4 factors:

a. For titrations by standard HAT, we used plasmas at 1/50 as the highest concentration. In other words, we used 2 μl of plasma per well, to react against 0.3 μl of RBCs. The plasma to RBC ratio was thus six times more than in HAT-field, where there are roughly equivalent volumes of RBCs and plasma, and this ratio of 1 cannot be altered since, in HAT-field, the blood samples are simply diluted before performing the assay. On this subject, we have found that increasing the amount of whole blood per well (in other words using blood that is less dilute) has very little influence over the HAT-field results, and, if anything, adding more blood can sometimes reduce the sensitivity, albeit never by more than 1 dilution.
b. Titrations for figure 6 were carried out on washed RBCs from an O-donor that were 5 days old, and, in our hands, RBCs which have been washed and stored at 4°C for a few days tend to work a bit better for HAT than those in whole blood.
c. When using washed RBCs, EDTA is no longer present, and we have found that excess EDTA can reduce HAT sensitivity for certain samples, possibly because the binding of certain antibodies may involve divalent cations. In the future, it may thus be interesting to explore the possibility of using heparin rather than EDTA as an anticoagulant.
d. Whilst plasma titrations were carried out by double dilutions of the plasmas, in the optimized protocol we have devised for HAT-field, the IH4-RBD reagent is titrated in steps of 3.16 fold, so as to cover a broader range (see Methods). This does, however, only really influence the upper right corner of the graph, i.e. the scores of samples with very high levels of antibodies.

On figure 6C, one finds four samples that scored positive by standard HAT, whilst the staining values in the Jurkat-S&R-flow test were in the doubtful zone between 10 and 40, and our view is that those samples probably contained some antibodies reacting specifically against the SARS-CoV-2 spike protein, and their more effective detection by hemagglutination than by FACS staining may be due to relatively high proportions of IgAs or IgMs. This is indeed reminiscent of the observation reported in our first paper that, during very early SARS-CoV-2 infections, HAT could detect antibody responses before CLIA (Townsend et al., 2021). Incidentally, although this is not something that we have yet managed to document formally, we noticed that the samples which contain sizeable amounts of IgMs and/or IgAs often reach higher HAT scores than expected from the FACS results obtained with the pan-human Ig secondary antibody, with even higher scores by plasma dilutions than in HAT-field (see data file). Of note, for sample 19, which was a false positive and gave very high titers against that IH4 alone, FACS analysis performed after the HAT assay showed that the human antibodies bound to the IH4-coated RBCs were predominantly IgMs (data not shown).

For one sample (n° 48 in data file), indicated by an orange symbol with a red circle in Figure 6C, the result of Jurkat-S&R-flow test was 44.84, i.e. just above the threshold value which is arbitrarily set to 40. This sample only led to partial hemagglutination at 1/50 in the standard HAT, but showed reactivity against the IH4 nanobody alone with an endpoint at 1/200 after centrifugation (score 3). We thus surmise that this sample does contained either very low or no specific anti-SARS-CoV-2 antibodies, which provides a justification for maintaining the Jurkat-S&R-flow test threshold at its current value.

As in all biological tests, increasing the sensitivity will almost unavoidably lead to an increase in the proportion of false positives. It is thus not surprising that, with the gain of sensitivity of HAT afforded by the combined use of PBN and spinning, the proportion of samples being detected as showing some reactivity against the IH4 nanobody should be higher than the 1 to 2% that were originally detected with the standard HAT protocol in various cohorts (Maurel Ribes et al., 2021; Townsend et al., 2021).

As can be seen on the table of data for the cohort which is provided as supplementary material, with the HAT-field protocol, after one hour under normal gravity, reactivity against the IH4 moiety was detected in 3% of samples (2 out of 60), and climbed to 8 % after spinning (5 out of 60). With the more sensitive protocol used for plasma titrations, 5 samples (8%) reacted against the IH4 nanobody after one hour under simple gravity, but this number climbed to 12 (20% of samples) after spinning. If the HAT-field test was ever to be used in a clinical context, the results would be invalidated for those samples found to react with the IH4 alone, and with such high frequencies as we have observed in our small cohort, this could be a significant problem. An alternative would be to perform, from the start, systematic parallel titrations of the IH4-BRD and IH4 alone reagent, in order to identify samples which react markedly better on IH4-RBD than on IH4 alone. This could be achieved either by using 20 μl of blood diluted into 600 μl to distribute in two sets of 8 wells, which would be quite easy since 20 μl corresponds to the second section on the plastic Pasteur pipets. Alternatively, titrations of the two reagents could be carried out over just 4 wells each, with larger dilution factors between wells (e.g. 10 fold).

In future, to reduce the proportion of false positives due to reactivities with the IH4 moiety, it may be interesting to investigate if the nanobody, which is of camel origin, could be somewhat “humanized” by site-directed mutagenesis without losing its capacity to bind to human glycophorin. Alternatively, the use of different antibodies binding to glycophorin, such as nanobodies derived from other species, or mAbs such as the one described by Kemp and colleagues (Kemp et al., 1988; Wilson et al., 1991) could also be explored as an alternative.

Two recent reports have described that HAT could be performed on cards rather than in V-shaped wells, with semi-quantitative results being obtained in minutes, which was made possible by the use of much higher amounts of the IH4-RBD reagent than when HAT is performed in V-shaped well (Kruse et al., 2021; Redecke et al., 2021). Reliably quantitative card-based tests HAT test would unquestionably be a very attractive solution for performing the test in field settings. Comparing the sensitivity, specificity and robustness of the two types of protocols, on cards or in V-shaped wells, on the very same cohorts of whole blood samples would be very interesting, especially if such comparisons were performed by third party laboratories.

## Conclusion

We have shown that the HAT-field approach, which uses a single drop of capillary blood, can provide, in a single step, a quantitative measurement of the antibodies against the viral RBD, which are those endowed with neutralizing activity. Such a test could prove very useful for identifying individuals in need of a vaccine boost (or a primary injection). Given its very low cost, the stability of the IH4-RBD reagent, and the simplicity of the procedure, HAT-field should be well suited to be performed pretty much anywhere, including in the poorest countries and the most remote corners of the globe. Given that HAT has already been successfully adapted to detect antibodies against the RBD of several SARS-CoV2 variants (Ertesvåg et al., 2021; Jeewandara et al., 2021a), we presume that it could be adapted very rapidly to evaluate the levels of antibodies reacting against the RBD of other newly arising SARS-CoV2 variants of concern, such as the newly arisen and very divergent Omicron variant, or, in future, to pretty much any new threatening pathogen, and could thus represent a great asset to be better prepared to face future pandemics.

## Methods

### Reagents

PBS and tissue culture media were all obtained from Gibco.

BSA Fraction V was obtained from Sigma (ref A8022 or A7888). Of note, we have tried using other sources of Fraction V BSA for preparing PBN, and found that they do not all work as well as the ones listed above to prevent veil formation in HAT.

Sodium azide (NaN3, Sigma S2002) was prepared as a 20% stock solution in milli-Q water and kept at room temperature. This 3M solution was then used as a 1000x stock for the preparation of PBN, PFN and PBS-azide.

PBN was prepared by adding 500 μl of the above 1000x Azide stock and 5 grams of BSA Fraction V (Sigma A8022) to 500 ml of PBS.

PFN, used for the dilution of antibodies and washes of FACS samples, was prepared by adding 500 μl of the above 1000x azide stock and 10 ml of fetal calf serum to 500 ml of PBS (we find that this is a very good use for unwanted or expired stocks of FCS that often clutter the bottom of freezers)

Because they contain azide (final concentration 3mM), no sterilizing filtration is needed for either PBN or PFN, and they can be kept for many weeks at 4°C.

Polyclonal anti-human Igs secondary antibodies, all conjugated to Alexa-488, were from Jackson laboratories, and purchased from Ozyme (France). Refs: anti-human Ig-GAM: 109-545-064, -G: 109-545-003, -A: 109-54-011, -M: 109-545-129

Anti RBD monoclonal antibodies: CR3022 (ter Meulen et al., 2006) and EY6A (Zhou et al., 2020) were obtained using antibody-expression plasmids, as previously described (Townsend et al., 2021).

Covid-19 sera 186, 197 and 203 were obtained from the virology department of the Toulouse hospital; plasmas 79 and 206 were from whole blood samples used in our previous paper describing the Jurkat-S&R-flow test (Maurel Ribes et al., 2021)

IH4-RBD (Wuhan and Delta) and IH4 alone were produced by transient transfection of HEK-293T cells, and purified from the supernatant by HIS-tag affinity purification, as previously described (Townsend et al., 2021). Highly concentrated stock solutions at 3 – 5 mg/ml in PBS were kept frozen as aliquots. Those were then used to prepare 100X stocks in either PBS or PBN, which were kept frozen, and kept a 4°C after thawing for up to a few weeks.

To study the stability of the IH4-RBD reagent over time, working solutions at 2 μg/ml in either PBS-azide or in PBN were prepared from 100X PBS stocks at various time points over the course of a whole year. Three aliquots of 500 μl each were then set aside from those working stocks, to be kept either at 4°C, at room temperature or at 37°C in a tissue culture incubator. The activity of those aliquots of working stocks was then evaluated at regular intervals by performing titrations of the IH4-RBD reagent against either the CR3022 monoclonal antibody diluted to a final concentration of 100 ng/ml, or various immune sera diluted to give similar endpoint to those obtained with CR3022.

When comparing the reactivities against the Wuhan and Delta IH4-RBD reagents, we ascertained that we were using working concentrations of the two reagents with comparable activities by performing titrations with two monoclonals, EY6A or CR3022, which recognize a binding site not affected by the two mutations carried by the RBD of the Delta viral lineage, and found the Wuhan and Delta reagents to have indistinguishable hemagglutinating activities under those conditions.

### Ethical statement

RBCs from O-blood donors were obtained from the Toulouse branch of the Etablissement Français du Sang (EFS), with whom the project was validated under agreement n° 21PLER2020-025.

Whole blood samples: The 60 samples used for figures 6, S1, S2 and S3 were routine care residues from patients of the Toulouse hospital, where all patients give, by default, their consent for any biological material left over to be used for research purposes after all the clinical tests requested by doctors have been duly completed.

According to the French law on ethics (loi Jardé), retrospective/prospective studies based on the exploitation of clinical care residues do not require to be submitted to an ethics committee. This study was reviewed and approved by the ‘Directeur de la Recherche et de l’Innovation du CHU de Toulouse’, who confirmed that this study conformed with all the ethical and legal requirements, and gave his signed approval with the agreement number RnIPH 2021-99. This was further confirmed by the signature of a Material Transfer Agreement between the Toulouse University Hospital and the CNRS, with the following references: Toulouse Hospital n° 20 427 C, CNRS n° 227232.

### Human samples

The 60 whole blood samples were collected, regardless of gender, in the course of the month September 2021. Those were anonymized within 24 hours of collection, transferred from the hospital to the research lab, and kept at room temperature until being used for HAT assays within 24 hours (i.e. less than 48 hours after blood samples were collected). In trial experiments, we had found that such samples could be stored for up to 5 days without any noticeable difference in the performance of the HAT tests.

After the whole blood samples had been used for HAT-field assays, the tubes were then spun, the plasmas harvested into fresh tubes, and sodium azide added to 3 mM final. Those harvested plasmas were kept at 4°C until they were used to perform the Jurkat-S&R-flow tests and HAT assays for plasma titrations.

The identities, clinical conditions and Covid status (PCR or positive serology) were unknown to the person performing the HAT experiments and Jurkat-S&R-flow tests.

Blood samples from O-donors (6 ml EDTA tubes) were obtained every few weeks from the EFS (Toulouse blood bank). Whilst whole blood is best kept at room temperature (i.e. between 20 and 25 °C), and can then be used for HAT assays for up to 5 or 6 days, washed red blood cells can be kept for several weeks, as long as they have been separated from the white blood cells, and are kept at 4°C in the right buffer (in our case Alsever’s solution, Sigma A3551).

For preparing RBCs for storage at 4°C, we used the following standard protocol. The EDTA collection tube is spun at 1000g for 20 minutes with no refrigeration, and the centrifuge brake set to 2 /9. The plasma is collected into a separate sterile tube and a total of 8 ml of sterile PBS used to resuspend the cells and transfer them to a 15 ml tube, on top of a 3 ml cushion of lymphocyte separation medium (Corning Ref 25-072-CV). This tube is then spun once more at 1000g for 20 minutes with no refrigeration, and the centrifuge brake set to 2/9. The supernatant, including the ring of white blood cells, is aspirated and discarded. The RBC pellet is then washed twice with 8 ml sterile PBS, once in 8 ml Alsever’s solution, before adding two volumes of Alsever’s solution to the one volume of packed RBCs. This tube of RBCs (at 30% v/v) can then be stored at 4°C, and used for HAT assays for several weeks. After a week of storage, however, we found that the RBCs progressively tend to lose their capacity to teardrop after spinning.

If whole blood from an O-donor was needed for experiments (e.g. as in Figure 2), 100-200 μl were transferred to a separate sterile tube before performing the above procedure, and this tube was kept at room temperature for a maximum of 4 days. Worthy of note, whilst all donors were seronegative at the start of this study (i.e. from summer 2020 to winter 2021, the proportion of seropositives started increasing in the spring 2021, correlating with the proportion of vaccinated people increasing in the French population, and we have come across no seronegative samples among the dozen of blood samples we have used since the beginning of the summer 2021. Whilst the serological status of the donors does not matter when working with washed RBCs, it would thus no longer be practical to plan performing HAT tests in the presence of 1% autologous plasma as originally recommended (Townsend et al., 2021), but, as shown in figure 1, this is no longer necessary in the presence of 1% BSA.

### Original HAT assays

For HAT assays performed under ‘standard’ original HAT conditions (Figures 1-4), outside of using PBN instead of PBS, we used similar reagent concentrations and incubation conditions to those defined in our original description of HAT (Townsend et al., 2021):

- in a final volume of 100 μl per well
- with approximately 0.3 μl of packed RBCs per well, i.e. 1 μl of 30 % stock stored in Alsever’s solution.
- using IH4-RBD at a final concentration of 1 μg/ml (i.e. 100 ng/well), or alternatively 0.5 μg/ml of IH4 alone since the size of the nanobody and the His tag corresponds to slightly less than half of that of the IH4-RBD recombinant protein.
- Taking pictures after incubating the plates for 60 minutes at room temperature, for which we find it very convenient to use a very simple home-made lightbox (https://youtu.be/e5zBYd19nIA).

For experiments such as that presented in Figure 1, tubes of the appropriate 2X stocks containing either the RBCs and the antibodies, or the IH4-RBD reagent were prepared to be mixed 50/50 in each well.

For titration experiments, we prepared stocks containing RBCs and the appropriate amount of the reagent to be kept constant (either the IH4-RBD reagent at 1 μg/ml or IH4 alone at 0.5 μg/ml for plasma titrations, or the appropriate antibody dilution for IH4-RBD titrations). 100 μl of those stocks were then distributed per well, and 200 μl for the wells of the first row. The reagents to be titrated were then added to each of the wells of the first row, and a multi-channel pipet was then used perform the serial dilutions by successive transfer of 100 μl to the wells of the adjacent row, with thorough mixing by pipetting gently up and down at least 6 times at each diluting step.

A similar method was used to perform the 2D titrations presented in figure 4, filling columns of wells with stocks of IH4-RBD serially diluted with the appropriate suspension of RBCs, and proceeding in a second stage to perform serial dilutions of the antibodies in rows, as described above.

### HAT-field assays

The optimized procedure we have arrived at and used for this study is based on using 7 serial dilutions of the IH4-RBD reagent. Rather than double dilutions, we elected to use 3.16 as a dilution factor between adjacent wells. This not only allows to cover a larger range of IH4-RBD concentrations, but because 3.16 is the square root of 10, two successive dilutions conveniently correspond to a factor of 10. The concentrations of the IH4-RBD stocks used to prefill the wells were thus, in ng/ml 4750, 1500, 475, 150, 47, 15, 4.7. After addition of one drop of diluted blood, i.e. roughly 30 μl, the final volume was thus ca. 90 μl, and the approximate final concentrations of IH4-RBD in the wells were thus, in ng/ml: 3160, 1000, 316, 100, 31, 10, 3.1.

A detailed step by step protocol on how to generate the stocks of those various dilutions, and how to perform the HAT-field test, is provided as supplementary material.

For every sample, the eighth well of a row is allocated to performing the very important negative control, which can consist of either PBN, or preferably PBN containing the IH4 alone, i.e. the nanobody without the RBD attached to it, at 1.6 μg/ml final concentration, corresponding to a molar concentration similar to that found in the well with the highest concentration of IH4-RBD.

For the 60 samples of the cohort used in our study, because we were running sets of parallel HAT-field assays with either the IH4-RBD-Wuhan or IH4-RBD-Delta reagents, we performed both types of negative controls by using PBN as a negative control in the plates for the Wuhan HAT-field assays, and IH4 alone as a negative control in the plates for the Delta HAT-field assays. As discussed above, we found that the negative controls using the IH4 alone were much more informative.

For spinning the plates at 100g for 1 min, we used a centrifuge with swinging-out trays (Eppendorf 5810R, set to 705 rpm)

### Scoring HAT

The procedure we used for scoring HAT tests was based on the same principle for plasma titrations using the original HAT protocol and for HAT-field: At the end of the incubation period, the plates were tilted to an almost vertical position using a purposefully designed homemade light box (see protocol provided, and [online video tutorial](https://www.youtube.com/watch?v=e5zBYd19nIA&feature=youtu.be)). The pictures used for scoring were taken with a mobile phone camera after about 20 seconds, i.e. when the teardrops in the negative control wells reached the bottom walls of the wells.

After transfer of the pictures to a computer, scoring was then carried out by simply counting the number of fully positive wells, starting either from the highest IH4-RBD concentration for HAT-field, or the least diluted plasma for titrations if using the original HAT protocol.

The trickiest part in scoring HAT lies with separating partially from fully positive wells. For this, we find that the most reliable means consists in considering any RBC pellet that shows a detectable pointy bottom as only partially positive (e.g. the fourth sample from the top on figure 7A).

**Figure 7:**
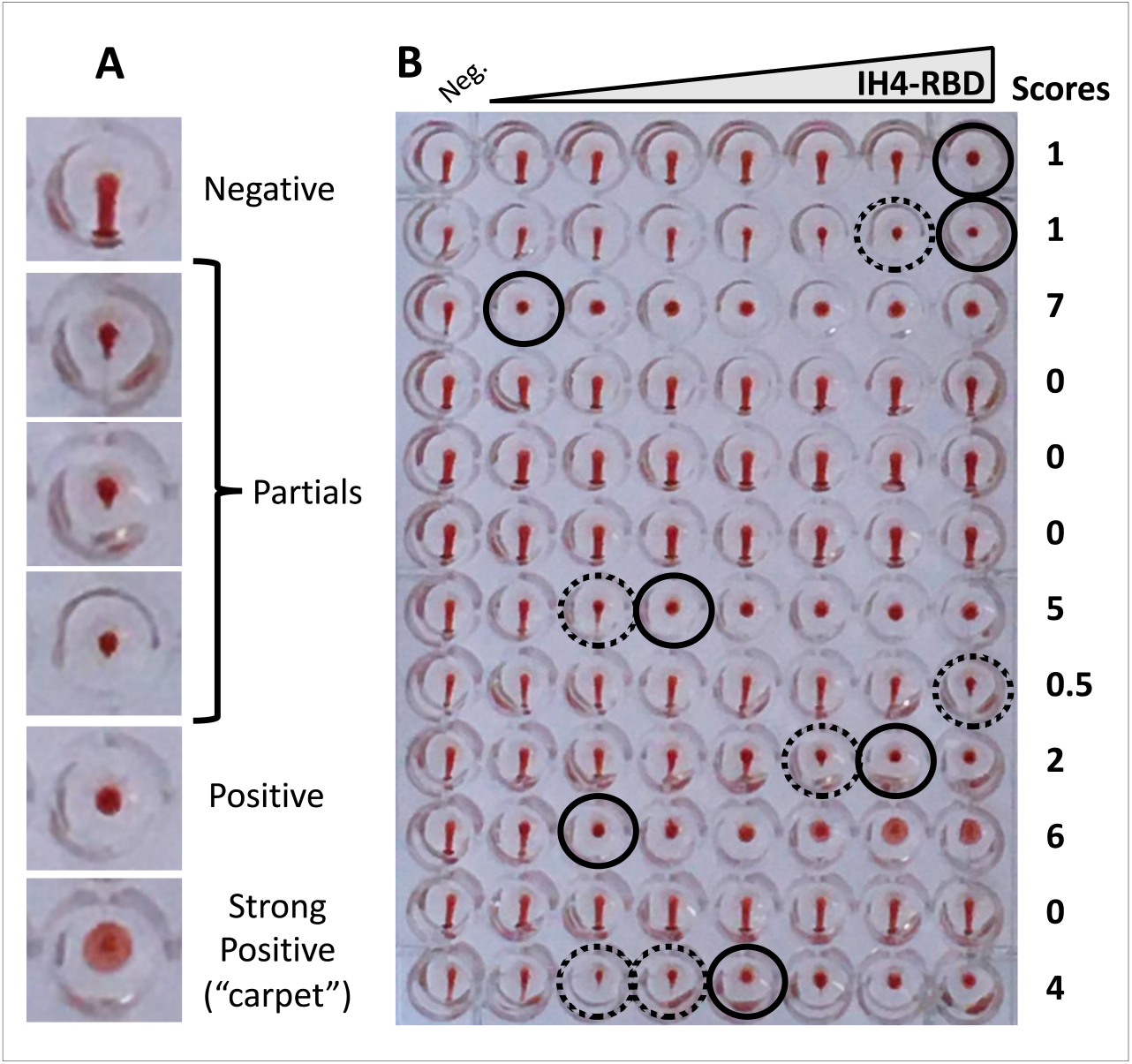
Scoring HAT. A: Examples of typical negative, partial and positive wells. Wells are considered as partial as soon as a small point appears at the bottom of the RBD pellet, such as that shown on the fourth line. B: Example of a plate with HAT-field performed with Wuhan IH4-RBD on 12 whole blood samples after 60 minutes under normal gravity. The final IH4-RBD concentrations in the wells from left to right are respectively 0, 3, 10, 31, 100, 316, 1000, 3160 ng/ml. Black circles indicate the last positive wells, and dotted circles partially positive ones. Scoring is carried out on the pictures by simply counting the number of positive wells, starting from the highest IH4-RBD concentration (i.e. from right to left).

With certain samples containing high amounts of antibodies, it is quite frequent to see the RBCs forming broader pellets, which we refer to as ‘carpets’ since those have a certain tendency to float off the bottom of the wells and fold upon themselves (see strong positive in Figure 7A, and sample on line 10 of 7B). Those carpets, which disappear upon centrifugation at 100g, are quite different from the veils due to the adsorption of IH4-RBD to the wells’ bottom in the absence of BSA (Figure 1A).

Whilst the tendency of certain samples to form carpets is highly reproducible, it is not seen for all samples with high levels of antibodies. For example, the sample on line 3 of figure 7B had even higher levels of antibodies than that on line 10, as determined both by HAT-field, plasma titration and FACS, but did not form carpets. Of note, the Ig-G/A/M profiles of samples as determined by FACS showed no obvious correlation with their tendency to form those carpets.

### Practical considerations for performing HAT assays

When performing HAT in PBN compared to PBS, we observed a slight increase in sensitivity which we suspect is most probably due to an improved sedimentation of the RBCs, with both azide, and BSA, contributing to the formation of more compact pellets at the bottom of the V-shaped wells. Regarding the role of azide, we postulate that, by blocking the metabolism of the RBCs, it probably increases their density and consequently their sedimentation. Regarding the beneficial role of facilitating the sedimentation of RBCs, this is something that Weggmann and Smithies had recognized soon after their initial description of the microtiter hemagglutination method, and for which they had proposed using paraffin-coated plates as an improvement (Wegmann and Smithies, 1968).

As we have seen, the improvement of HAT sensitivity provided by prolonging incubations up to 5 hours can be very advantageously replaced by a brief step of centrifugation at 100g, which can be performed after only 15 minutes of incubation. The need for a centrifuge with the capacity to spin 96 well plates would, however, rather preclude the possibility of performing HAT in the field. But accelerations of 100g are in the range of those attained by hand-driven centrifuges such as [salad spinners](https://www.youtube.com/watch?v=797-uBf8A1Q). We have investigated the possibility of using this type of centrifuge for spinning 96 well plates after just 15 to 20 minutes of incubation, but have found that this only works for the central 2 columns of a 96 well plate because the RBCs in the outer columns are pushed to the outside of the wells. Whilst it is hard to conceive that hand-driven centrifuges with the capacity to spin 96 well plates in swinging-out trays could become part of the equipment enabling the use of HAT in the field, a rather simple solution would be to design plastic strips of conical wells for individual tests. Using very simple adapters, those purposefully designed sets of wells could then be spun in hand-driven centrifuges of the salad spinner type. One aspect that will have to be considered for the design and use of such individual strips of wells will be to ensure that, upon storage, the various dilutions of IH4-RBD are as stable in such strips as the working stocks of IH4-RBD (2 μg/ml) tested in Figure 3. The design, and manufacture, of such disposable strips of wells, which would necessarily require the involvement of an industrial partner, was, however, well beyond the means of this study.

One important consideration about performing centrifugation at the end of a HAT assay is that, whilst this works very well with RBCs contained in whole blood samples, or with washed RBCs from freshly collected blood, we found that it can become problematic with RBCs that have been stored at 4°C for more than a week. Over time, stored RBCs will indeed progressively lose their capacity to teardrop, which can, incidentally, result in a slight improvement of the apparent sensitivity of HAT assays performed under simple gravity. But, if submitted to accelerations of 100g, we have found that such “aged” RBCs will form compact pellets that will fail to teardrop, even upon prolonged tilting of the plate. (of note, this is also the case for RBCs that have been kept in azide for a few hours. Preparing suspensions of RBCs in PBN should thus be done just before performing the HAT assays).

In their original paper, Wegmann and Smithies had described using incubations of 4 to 6 hours as their standard protocol, and suggested that a brief step of centrifugation could be used as an alternative (Wegmann and Smithies, 1966). For the record, during the initial stages of this study, we had made several attempts to use centrifugation, but we did not have access to whole blood samples at the time. We were thus using O-RBCs which had often been stored at 4°C for up to several weeks, and had initially given up on centrifugation because the aged RBCs we were using failed to teardrop after centrifugation. Incidentally, in that same paper, Wegmann and Smithies also suggested using BSA as an alternative to serum or plasma to promote better RBC settling patterns. They were, however, recommending using BSA at 0.22% which we found to be less effective than 1% to block the adsorption of the IH4-RBD reagent to plastic.

### Jurkat-S&R-flow test

Briefly, Jurkat-S and Jurkat-R cell lines, obtained and grown as previously described (Maurel Ribes et al., 2021), were resuspended in their own tissue culture medium at a concentration of 2.2 10^6^ cells/ mL before pooling equal volumes of the two.

Plasmas to be tested were diluted 1/10 in PFN (PBS / 2% FCS / 200 mg/L sodium azide). 20 μL of these 1/10 dilutions were then placed in U-bottom 96 well plates, before adding 180 μl per well of the Jurkat-S&R mix.

The plates were then incubated for 30 minutes at room temperature before placing them on ice for a further 30 minutes. All subsequent steps were carried out in the cold, with plates and washing buffers kept on ice. After the primary staining, samples were washed in PFN, with resuspending the cells by tapping the plate after each centrifugation, before adding 150 μl of PFN for the next wash. After 2 washes, the samples were split into 4 wells, and all resulting samples were washed one last time.

One drop (i.e. ca. 30 μl) of either the pan-specific anti-Ig-GAM secondary fluorescent antibodies, as well as anti-IgG, -IgA or -IgM, all diluted 1/200 in PFN was added to each of the four wells for each sample, and the cells resuspended by gentle shaking of the plates. After an incubation of 60 min on ice, samples were washed two more times with cold PFN before transferring the samples to acquisition tubes in a final volume of 300 μl PFN containing 30 nM TO-PRO™-3 Iodide (Thermo Fischer Scientific, ref T3605).

The samples were then analyzed on a FACScalibur flow cytometer controlled by the Cellquest pro software (Version 5.2, Beckton Dickinson), using the FL1 channel for Alexa-488, the FL3 channel for m-Cherry, and the FL4 channel (with the 633 nm laser) for live gating with the TO-PRO™-3 live stain. Post-acquisition analysis of all the samples was performed using the Flowjo software (version 10.7.1). The values used as results are those for specific staining, i.e. the difference between the GMFI (geometric mean fluorescent index) measured on the Jurkat cells expressing the SARS-CoV-2 spike protein and the control Jurkat cells expressing the mCherry fluorescent protein. The value of 40 of specific staining was used as the threshold above which the samples were considered as positive. As described previously, with the cytometer settings used in this study, this correspond to 20 fold the value obtained with cells stained just with the secondary antibody (Maurel Ribes et al., 2021).

### FACS analysis of RBCs after HAT

To quantify the amount of antibodies bound to the RBCs’ surface after a HAT assay, an adjustable pipet was used to resuspend the RBCS by pipetting up and down several times, and 15 μl (out of 90 or 100) were transferred to the well of a U-bottom 96 well plate pre-filled with 150 μl PFN. The RBCs were then washed by three repeated sequences of centrifugation at 800 g for 3 min, followed by flicking the supernatant out, tapping the plate, and adding 150 μl of PFN. One drop (i.e. ca. 30 μl) of anti-human secondary antibody conjugated to alexa-488, diluted 1/200 in PFN was added to each of the wells, and the cells resuspended by gentle shaking of the plates. After an incubation of 60 min on ice, samples were washed two more times with cold PFN before transferring the samples to acquisition tubes in a final volume of 300 μl PFN. The samples were then analyzed on a FACScalibur flow cytometer controlled by the Cellquest pro software (Version 5.2, Beckton Dickinson). Post-acquisition analysis of all the samples was performed using the Flowjo software (version 10.7.1)

Because they can be obtained in very large numbers and stored at 4°C for several weeks, it is actually much simpler to use RBCs for FACS analysis than Jurkat cells, which need to be kept in culture continuously. But we find that FACS analysis of RBCs has a much reduced dynamic range compared to the Jurkat-S&R-flow test. It is indeed less sensitive, and many samples harboring low levels of antibodies would not be detected by RBC staining. And for samples that contain very high amounts of antibodies, the RBCs will tend to stay agglutinated, and the resulting cell clumps will be discounted during FACS analyses, with samples containing much lower numbers of usable cells, and the FACS results skewed towards lower values. If wanting to perform FACS analysis of RBCs after HAT, a solution to avoid this problem of clumping is to analyze those samples that are just one or two dilutions above the agglutination endpoint. But this will mean that all samples will not all have been stained with the same amounts of reagent and consequently that the staining levels cannot be compared with one another.

Alternatively, the problem can also be avoided by keeping the concentrations of the IH4-RBD below 100 ng/ml, but this will result in a further reduction of the sensitivity for the samples with low levels of antibodies.

## Supporting information

Data cohort clinical samples

## Data Availability

All data produced in the present study are available upon reasonable request to the authors

## Author contributions

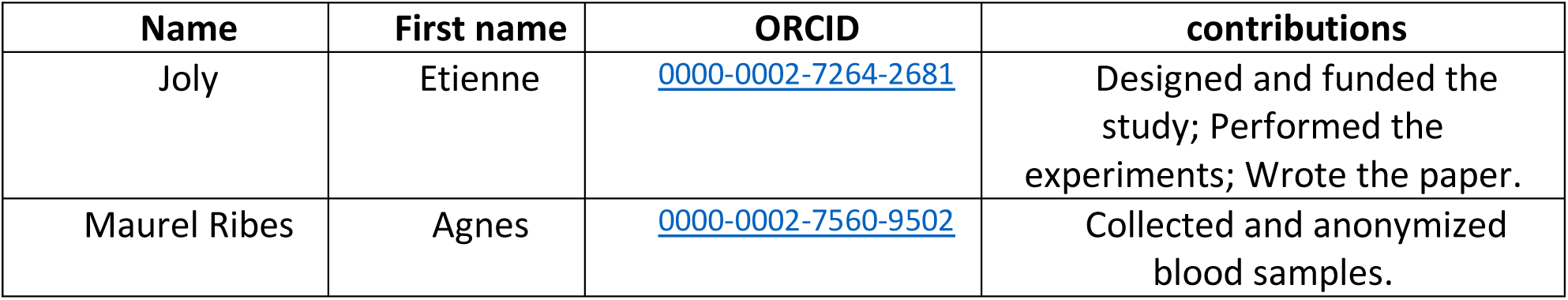

## Other contributors

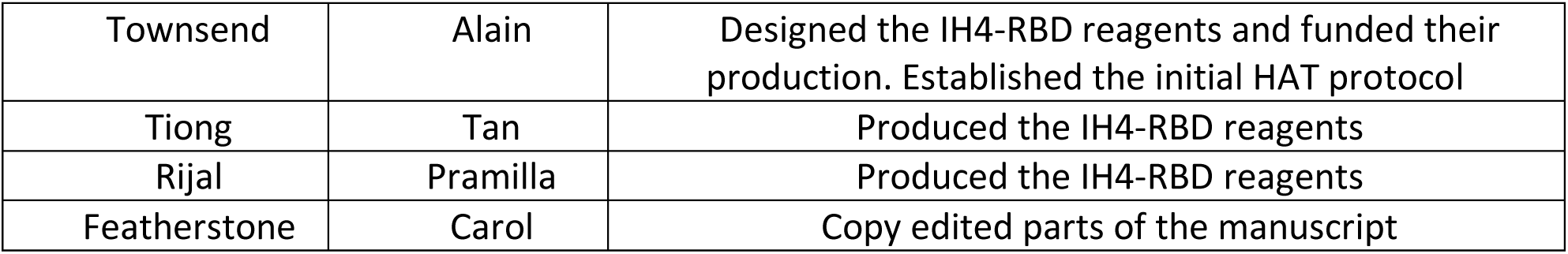

|  |  |  |
| --- | --- | --- |
| Townsend | Alain | Designed the IH4-RBD reagents and funded their production. Established the initial HAT protocol |
| Tiong | Tan | Produced the IH4-RBD reagents |
| Rijal | Pramilla | Produced the IH4-RBD reagents |
| Featherstone | Carol | Copy edited parts of the manuscript |

## Acknowledgements

We are particularly grateful to the following people: Alain Townsend, Tiong Kit Tan, Pramila Rijal, who contributed the various materials detailed in the above table, and to Carol Featherstone for copy editing. We also gratefully acknowledge the contributions of Marianne Navarra, for her help in setting up the agreement with the hospital; Emmanuelle Naser and Pénélope Viana from the IPBS flow cytometry facility for their assistance; the staff of the Toulouse EFS (Etablissement Français du Sang) for the provision of O-blood samples; Florence Abravanel and Jacques Izopet for providing immune sera from convalescent Covid patients, and Eric Vallée for the experiments he carried out very early on in the course of the pandemic to compare the performance of HAT with those of the lateral flow assays.

## Supplementary material

### 1) Data file for the clinical samples

With the cohort of 60 clinical samples of whole blood we used to validate the performance of the modified HAT protocols, we ended up producing 24 different sets of data. Whilst a good number of different comparisons between those sets are presented on figures 6 and S1-3, it was not practical to provide comparisons for all the possibly interesting combinations. With the sets of data provided as an Excel file, interested reader wanting to explore other comparisons not provided here can easily perform those.

The data sets provided are as follows:

| Column |  |
| --- | --- |
| A | Sample numbers |
| **HAT-field scores** (starting with IH4-RBD at 3.16 µg/ml)<br>Each score increment corresponds to dilution of IH4-RBD by a factor of 3.16 |  |
| B | IH4-RBD Wuhan after 60' under normal gravity |
| C | IH4-RBD Wuhan after 60' + 1' spin @ 100g |
| D | IH4-RBD Delta after 60' under normal gravity |
| E | IH4-RBD Delta after 60' + 1' spin @ 100g |
| F | IH4-RBD Wuhan 1' spin @ 100g after 15' incubation |
| G | IH4-RBD Delta 1' spin @ 100g after 15' incubation |
| H | IH4-RBD Wuhan after 1 hour under normal gravity |
| I | IH4-RBD Wuhan after 3 hours under normal gravity |
| J | IH4-RBD Wuhan after 5 hours under normal gravity |
| K | IH4-RBD Delta after 1 hour under normal gravity |
| L | IH4-RBD Delta after 3 hours under normal gravity |
| M | IH4-RBD Delta after 5 hours under normal gravity |
| **Titration on O- RBCs** (using 1 µg/ml IH4-RBD)<br>Starting with plasmas @1/50 (score 1)<br>Each score increment corresponds to one further double dilution of plasma |  |
| N | IH4-RBD Wuhan after 60' under normal gravity |
| O | IH4-RBD Wuhan after 60' + 1' spin @ 100g |
| P | IH4-RBD Delta after 60' under normal gravity |
| Q | IH4-RBD Delta after 60' + 1' spin @ 100g |
| **Reactivities on IH4 alone** (performed only on those samples which had shown reactivity on IH4 alone @ 1.6 µg/ml ) in the initial whole blood screen |  |
| **HAT-field scores** (starting with IH4 alone at 1.6 µg/ml)<br>Each score increment corresponds to dilution of IH4 alone by a factor of 3.16 |  |
| R | IH4 alone after 60' under normal gravity |
| S | IH4 alone after 60' under normal gravity + 1' spin @ 100g |
| **Titration on O- RBCs** (using 0.5 µg/ml IH4 alone)<br>Starting with plasmas @1/50 (score 1)<br>Each score increment corresponds to one further double dilution of plasma |  |
| T | IH4 alone after 60' under normal gravity |
| U | IH4 alone after 60' under normal gravity + 1' spin @ 100g |
| **Results of the Jurkat-S&R-flow test** (GMFI Jurkat-S – GMFI Jurkat-R ) |  |
| V | All human Igs: ( red scale was applied to symbolize signal intensities ) |
| W | IgG ( blue scale was applied to symbolize signal intensities ) |
| X | IgA ( yellow scale was applied to symbolize signal intensities ) |
| Y | IgM ( green scale was applied to symbolize signal intensities ) |
| Comparison of the signals obtained for IgG / IgA / IgM (bar for negative samples) |  |
| Z | Sums of columns W (IgG) + X ( IgA) + Y (IgM) |
| AA | Percentage of signal for IgG (blue scale) |
| AB | Percentage of signal for IgA (yellow scale) |
| AC | Percentage of signal for IgM (green scale) |
| **Clinical information** |  |
| AD | Gender of patient |
| AE | Covid / PCR History |

### 3) Step by step protocol for HAT-field

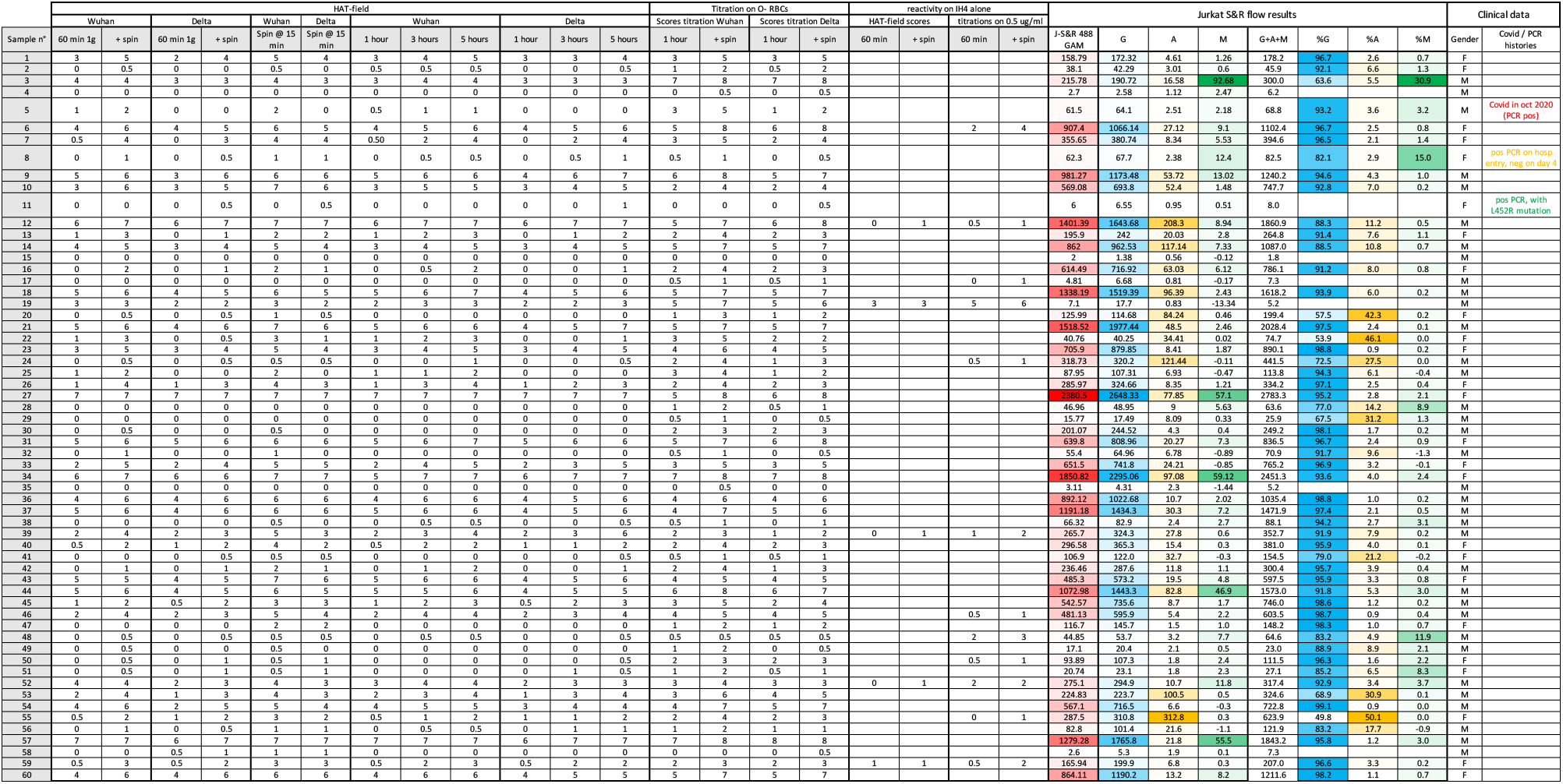

## Supplementary figures

**Supplementary figure 1:**
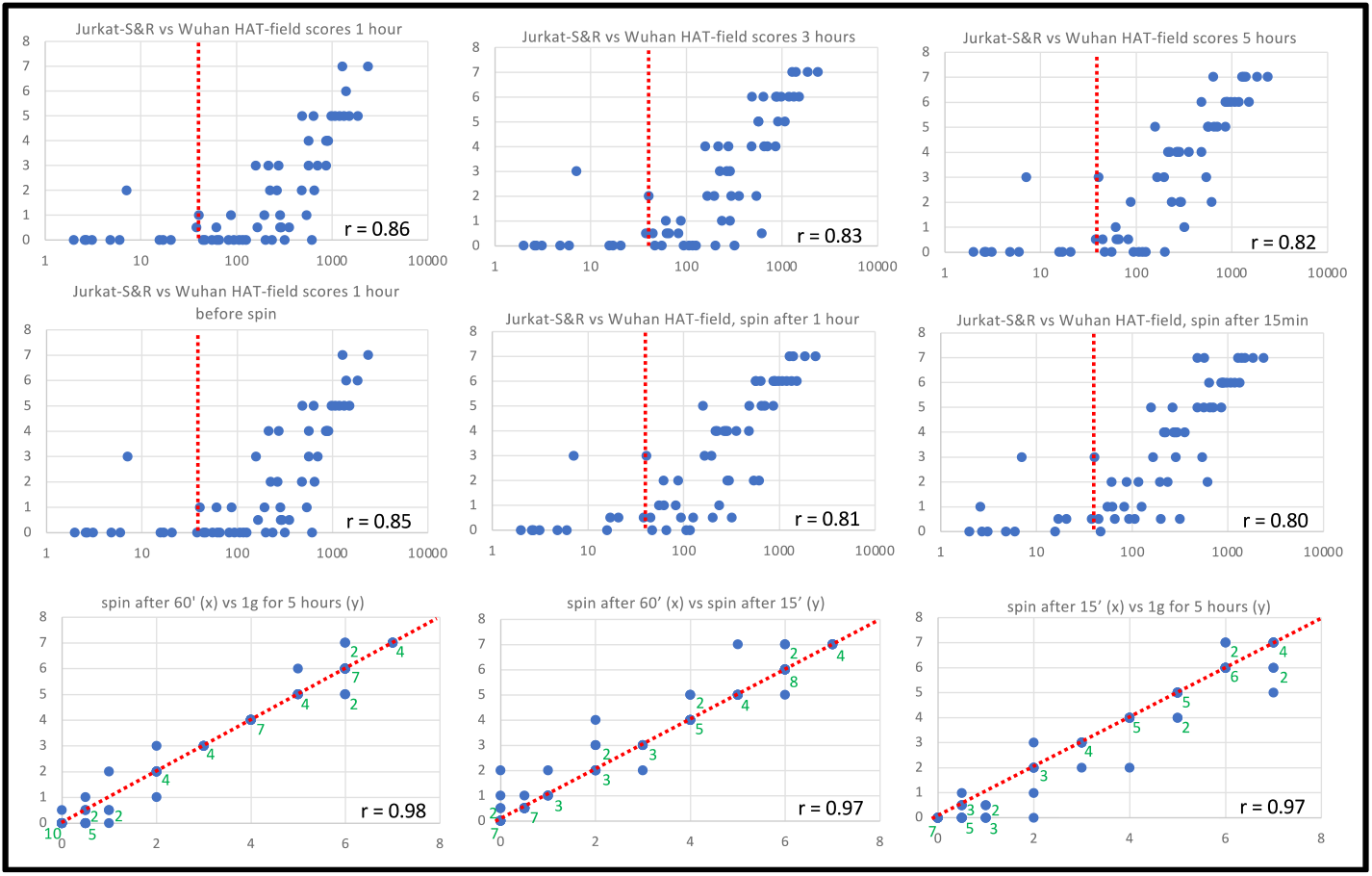
Influence of the incubation times on HAT-field scores For each of the 60 samples of the cohort of clinical whole blood samples, HAT-field was setup in three separate parallel plates, as described in Methods. The three separate plates were then submitted to the following incubation conditions:

i. Under normal gravity (1g), with tilting and taking pictures at 1, 3 and 5 hours before returning the plates to a horizontal position each time.
ii. under normal gravity for one hour, then taking pictures before and after spinning the plates for 1 minute at 100g
iii. incubating the plates for just 15 minutes horizontally at 1g before spinning it for 1 minute at 100g and then taking the pictures. This resulted in 6 data sets, which were all compared to the values of the Jurkat-S&R-flow test (first two lines). The vertical doted red lines indicate the threshold for positive samples used for the Jurkat-S&R-flow test. For the HAT-field test, although we recorded the partially hemagglutinated wells at the highest IH4-RBD concentration as a value of 0.5, we only consider the fully hemagglutinated wells as truly positive (score 1). Comparison of the 3 graphs on the first line shows that, when the plates are incubated under normal gravity, the sensitivity of the detection increases with incubation times, with fewer and fewer samples identified as positives by the Jurkat-S&R-flow test (i.e. past the red line) with HAT-field scores of zero or 0.5. The first graph on the second line is effectively a repeat of the one just above, i.e. 1g for 60 minutes. Comparison of the two shows that, although there are minor differences (i.e. the HAT-field scores of a few samples differing by just one unit), the method is highly reproducible. The next two graphs correspond to the results obtained after spinning the plates at 100g for 1 minute after either 60 minutes (2^nd^ graph), or after just 15 minutes (3^rd^ graph), giving results that are very comparable to one another, and to those obtained after incubating the plates for 5 hours under normal gravity. The graphs on the third line are one on one comparisons between the sets of scores obtained under these three latter conditions, i.e. 1g for 1 hour then spin, 1g for 5 hours, or 1g for just 15 minutes before spinning. On this series of graphs, because of the discrete nature of the scales used for both the X and Y axes, many points overlap on top of one another. In such cases, their numbers are indicated by the adjacent green numbers. Although the results obtained with those three conditions are very closely related to one another, it is worthy of note that spinning the plates after just 15 minutes does result in a few samples with slightly higher scores than those obtained with the other two conditions.

**Supplementary figure 2:**
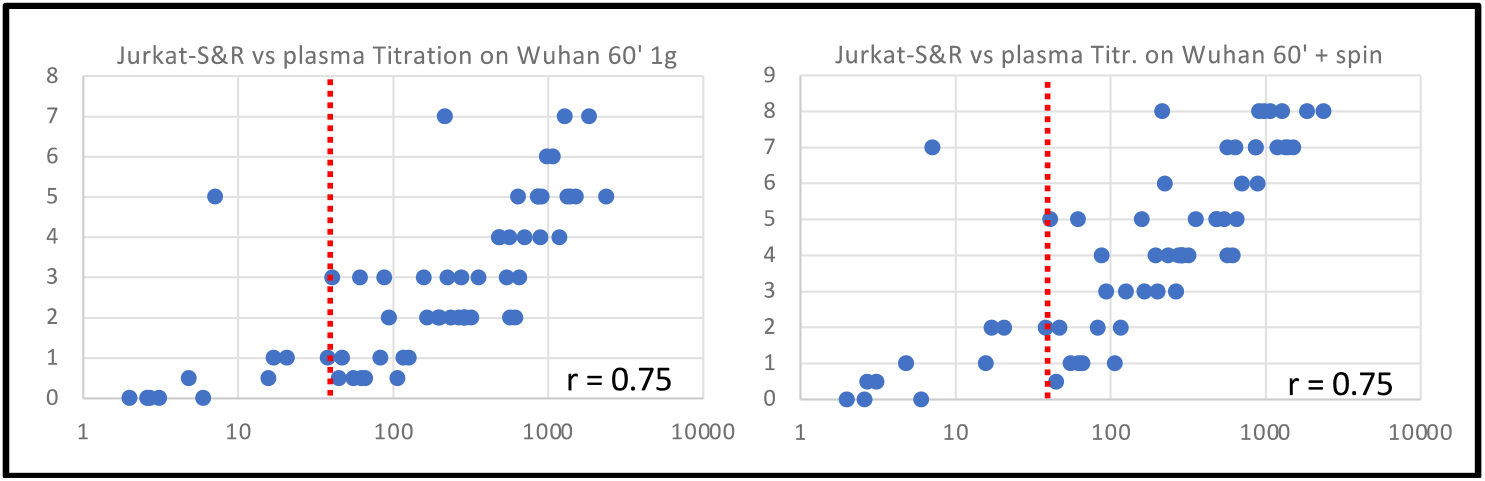
Spinning of plates for plasma titrations after 60 minutes incubation under normal gravity also greatly increases sensitivity. As can be seen by comparing the two graphs below, when performing plasma titrations using the standard HAT protocol, spinning the plate after 60 minutes of incubation at 1g (right panel) results in a marked improvement of the sensitivity, with all of the samples that were detected as positive by FACS having a HAT score of at least 1, i.e. showing full hemagglutination at 1/40, the highest plasma concentration used. The one sample which gave partial hemagglutination and FACS signal just above the positive threshold was actually most probably a false positive (see figure 6 and sample 48 in data file). See Discussion for considerations relating to the importance of using the IH4 alone reagent as a negative control to counter the decrease in specificity linked to increased sensitivity.

**Supplementary figure 3:**
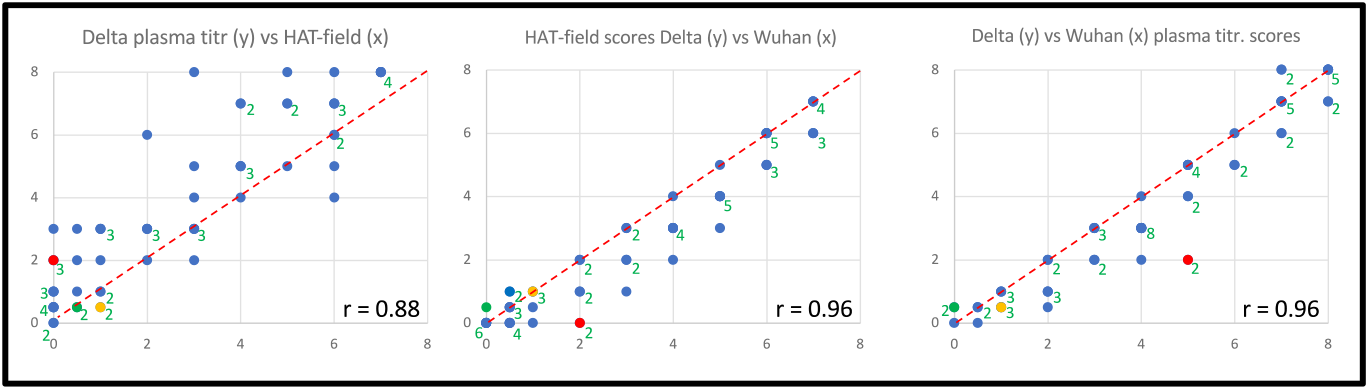
Comparing scores obtained with IH4-RBD-delta to those obtained with IH4-RBD Wuhan The above graphs were generated using: For HAT-field, the scores obtained after spinning the plates at 100g for 1’ after 15’ incubation at 1g (columns F for Wuhan and G for Delta in the data file). For the plasma titrations, the scores obtained after spinning the plates at 100g for 1’ after 60’ incubation at 1g (columns O for Wuhan and Q for Delta in the data file). When several points overlap, their numbers are indicated by the adjacent green numbers. The first graph shows that, as was seen in figure 6A with the IH4-RBD Wuhan reagent, under the conditions used in this study, plasma titrations are, overall, more sensitive than HAT-field. The second and third graph show that, for both HAT-field and plasma titrations, the scores obtained with the IH4-RBD Delta reagent tend to be similar or lower by one or two units than those with the IH4-RBD Wuhan reagent. The validation of the optimized HAT protocols was performed on a cohort of clinical whole blood samples collected in the course of the month of September 2021. At that time, most of the SARS-CoV-2 virus circulating in France belonged to the Delta lineage. Since those were blood samples from hospitalized patients, we were expecting to find a sizeable proportion of Delta-infected patients. Somewhat disappointingly, gathering of clinical information on the patients from which the blood samples were obtained, which took place after the experiments were carried out, actually revealed that, in our cohort of 60 samples, only 3 were from patients who had ever had a positive PCR for SARS-CoV-2. The first patient (sample 5, red spots) had had a positive PCR in October 2020, and must thus have been infected by a viral strain different from Delta. The level of the plasmatic anti-spike antibodies measured by the Jurkat-S&R-flow test was low (specific staining signal of 61.5), dominantly of the IgG isotype (93.2 %), and higher HAT scores with the Wuhan IH4-RBD than with the Delta IH4-RBD: HAT-field : W=2, D=0 ; HAT titration: W= 5, D=2 The second patient (sample 8, yellow spots) had a positive PCR on admission, which was the day when the blood sample we used was collected, but no information on viral genotyping was available. A PCR carried out 4 days later was negative. The level of the plasmatic anti-spike antibodies measured by the Jurkat-S&R-flow test was also low (specific staining signal of 62.3), with a sizeable signal for IgMs (15%). HAT scores were only just positive: HAT-field : W=1, D=1; HAT titration: W= 1, D=0.5. Based on this limited amount of information, no conclusion can be drawn regarding the lineage of the virus that infected that patient. The third patient (sample 11, green spots) had a positive PCR on admission, which revealed a viral strain carrying the L452R corresponding to the delta lineage. No further information is available about the evolution of the infection. Plasmatic anti-spike antibodies were undetectable with the Jurkat-S&R-flow test (specific staining signal of 6). Among the 16 sets of HAT scores (columns B-Q), although none were clearly positive, three were recorded as partially positive, all with the IH4-RBD Delta reagent : HAT-field 60’ + spin and 15’ + spin, and titration 60’ + spin. Although the data obtained for those three samples can only be considered as anecdotal, it is nonetheless comforting to see that they all follow the expected direction. A properly setup clinical trial, enrolling larger numbers of patients infected by various strains of the SARS-CoV-2 virus would need to be setup to formally validate the performance of the two reagents, and their capacity to follow serological responses after infection with those various strains. This was, however, far beyond the reach of this study.

## Protocol for HAT-field

### Equipment Required

#### Equipment for laboratory procedures

- Adjustable pipets + tips
- 15 ml tubes and 10 ml pipets
- Small tubes (e.g. Eppendorf tubes)
- V bottom 96 well plate (e.g. Sarstedt 82.1583)

### Reagents

- **RBC dilution** = PBS + 2 mM EDTA
- **PBN** = PBS + 1% BSA + 200 mg/l azide
- **100 X Stock of IH4 alone** @ 240 μg/ml in PBN
- **100 X Stock of IH4-RBD** @ 475 μg/ml in PBN
- Control Antibody CR3022 @ 20 ng/ml in PBN (other mAbs, e.g. EY6A can also be used)

### Equipment for HAT-field procedure

- Disposable lancets (e.g. Sarstedt 85.1016), sterile swabs, alcohol
- Small tubes (e.g. Eppendorf tubes) containing 300 μl of RBC dilution (PBS + 2 mM EDTA)
- Plastic Pasteur pipettes (small volume e.g. Sarstedt 86.1180)
- V bottom 96 well plates, if possible prefilled with 60 μl/well of working solutions (see below).

### Preparation of working solutions

Rather than simple DD, we elected to use a factor of 3.16 between successive dilutions. This not only allows to cover a larger range of IH4-RBD concentrations, but because 3.16 is the square root of 10, two successive dilutions correspond to a factor of 10. The concentrations of the IH4-RBD stocks used to prefill the wells will thus be, in ng/ml 4750, 1500, 475, 150, 47, 15, 4.7. After addition of one drop of diluted blood, i.e. roughly 30 μl, the approximate final concentrations of IH4-RBD in the wells will thus be, in ng/ml: 3160, 1000, 316, 100, 31, 10, 3.

Label eight 15 ml tubes: IH4 Control, and 1 to 7

Tube IH4 Control, will contain 10 ml PBN + 100 μl **100 X Stock of IH4 alone** @ 240 μg/ml in PBN To perform serial dilutions of the IH4-RBD working solutions

1. Place 14.85 ml PBN in tube n°1, and 10.25 in all others.
2. Add 150 μl of 100 X IH4-RBD Stock to Tube n°1 > 4.75 μg/ml IH4-RBD
3. Use 10 ml pipet to transfer 4.75 ml to tube n°2 > 1.5 μg/ml IH4-RBD
4. Use the same 10 ml pipet to transfer 4.75 ml to tube n°3 > 470 ng/ml IH4-RBD
5. Use the same 10 ml pipet to transfer 4.75 ml to tube n°4 > 150 ng/ml IH4-RBD
6. Use the same 10 ml pipet to transfer 4.75 ml to tube n°5 > 47 ng/ml IH4-RBD
7. Use the same 10 ml pipet to transfer 4.75 ml to tube n°6 > 15 ng/ml IH4-RBD.

7) Use the same 10 ml pipet to transfer 4.75 ml to tube n°7 > 4.7 ng/ml IH4-RBD.

NB: Tube n°7 will contain 15 mls whilst the others will contain just 10.25 ml.

Once prepared, the working solutions can be kept for several weeks, or even months, at either 4°C or room temperature without any noticeable loss of activity.

### HAT-field Procedure

#### Plate preparation

If an adjustable pipet is available, use it to prefill the wells of a V-bottom 96-well plate with 60 μl/well of the various working solutions (Column 1: IH4 negative control, Columns 2-8 : IH4-RBD dilutions 1 to 7)

If out in the field with no access to an adjustable pipet, the reagent can be distributed just before the assay will be performed, using a plastic Pasteur pipet to dispense 2 drops (i.e. ca. 60 μl) of the working solutions in the appropriate wells, and working in increasing order of concentrations (IH4-RBD dilutions 1 to 7), and a different pipet for the IH4 nanobody negative control.

<u>Blood collection and assay</u> (Subject should be asked to clean hands with soap, using warm water if possible, and to dry them well so that the blood drop stays compact on the fingertip).

- Take a plastic Pasteur pipet, and make sure that you have spotted the limit between the first and second section, which corresponds to 10 μl. Take one of the small tubes with 300 μl of PBS-EDTA, and label it with the subject’s name.
- Wipe finger pulp with sterile towel/swab. Prick skin on the outside of finger pulp with disposable, single use lancet. Wipe away first drop with same sterile swab
- Massage second drop of blood to the size of a lentil
- Take up 10 μl with the Pasteur pipet (first section filled), and transfer this blood to the small tube containing 300 μl of PBS-EDTA. Pipet up and down a couple of times, avoiding making bubbles.
- Using the same Pasteur pipet, take up all 310 μl of diluted blood, and distribute one drop into each of the 8 adjacent wells of a row, containing the IH4 control and IH4-RBD dilutions 1 to 7.
- Incubate the plate in a horizontal position for a minimum of 1hr at ambiant temperature. The red blood cells will sediment and form red dots at the bottom of the wells. Alternatively, if a plate centrifuge is available, plates can be spun at 100g for 1 minute after just 15 minutes incubation.
- Tilt plate at ca. 10° from the vertical against a well-lit white background (if possible on a lightbox, see below)
- When the RBCs have formed teardrops that reach all the way to the walls of the control wells (this should normally take less than 30 seconds), take photographs of the plate to record the results (with a smartphone, for example), making sure the camera is at least 50 cms away from the plate so that the bottom of all the wells can be clearly seen in the pictures. For this, we find that a very simple lightbox can greatly improve the ease of tilting the plates, and the quality of the pictures (see tutorial on https://youtu.be/e5zBYd19nIA)
- Scoring of the samples is done by counting the wells from the highest IH4-RBD concentration to the last well showing complete hemagglutination, i.e. a symmetrical dot with no “pointy bottom” as a result of the tilting.

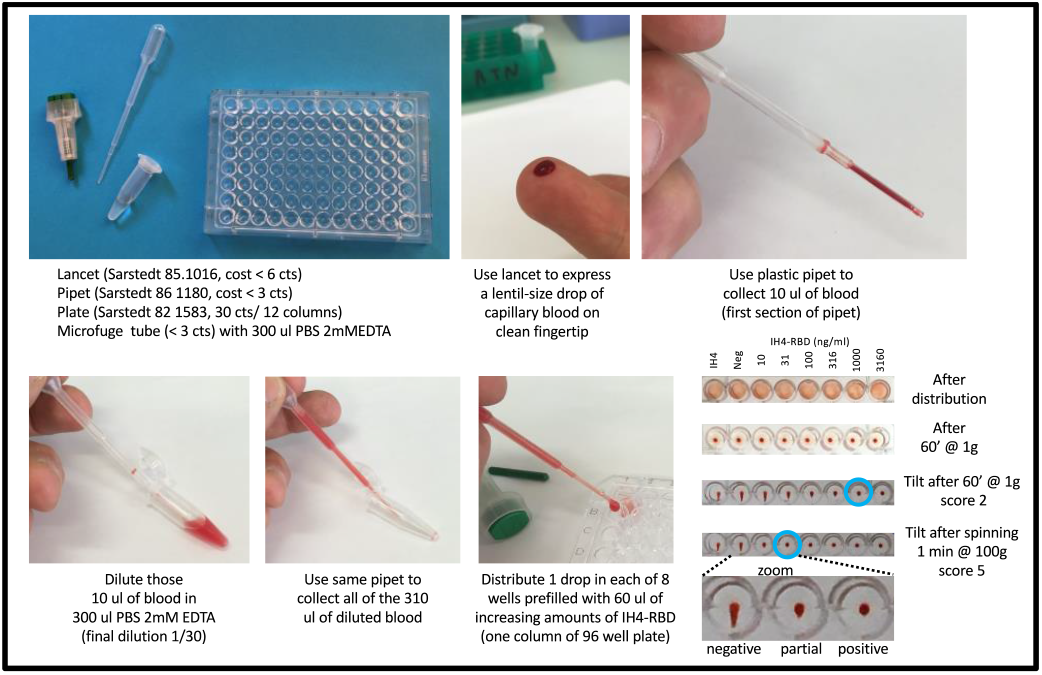

### Remarks

When performing the assay for the first time, once the 10 μl of blood have been diluted in the 300 μl PBS-EDTA, it is a good idea to start by counting the drops you will obtain by dropping them back into the tube used for the dilution, so as to ensure that you can obtain 8 drops (each drop should be just under 30 μl). If you do not, try placing the pipet at a different angle to generate smaller drops.

If a plate centrifuge is not available, longer incubations, up to 5 hours, will results in increased sensitivity. To this end, plates can be photographed after one hour, then returned to the horizontal position, and the tilting/photographing repeated at 3 and 5 hours (or other times if more convenient).

Rather than PBN, the negative control should preferably consist of the IH4 nanobody at a final concentration of 1.5 μg/ml (i.e. corresponding to the molar concentration of the IH4-RBD reagent in tube 1). Samples that show reactivity against IH4 will then have to be excluded.

Alternatively, a second row of wells containing titrations of the IH4 nanobody alone can be used to assess the plasmatic reactivity against the IH4 moiety of the IH4-RBD reagent. For this, collect 20 μl of blood by filling the plastic Pasteur pipet to the second section, and dilute this blood in 600 μl of PBS-EDTA, before using the pipet to place one drop in the 16 wells of the two rows.

For better quality photographs, it is best to use a lightbox. A very simple and cheap lightbox can be made with a cardboard box and a small lamp, such as the examples below, and in the tutorial video found at the end of this link: https://youtu.be/e5zBYd19nIA

If there are no positive samples in the set tested, the activity of the IH4-RBD can be ascertained by using a plastic Pasteur pipet to add one drop of the CR3022 mAb at 20 ng/ml to all the wells of one row. After returning the rest of the CR3022 stock to its tube, that same pipet can then be used to resuspend the RBCs in all the wells of that row, starting by the well with the lowest concentration of IH4-RBD and working towards higher concentrations, avoiding the formation of bubbles as much as possible. After a further hour of incubation, if stocks of IH4-RBD and CR3022 are fully functional, the endpoint should be around a score of 5.

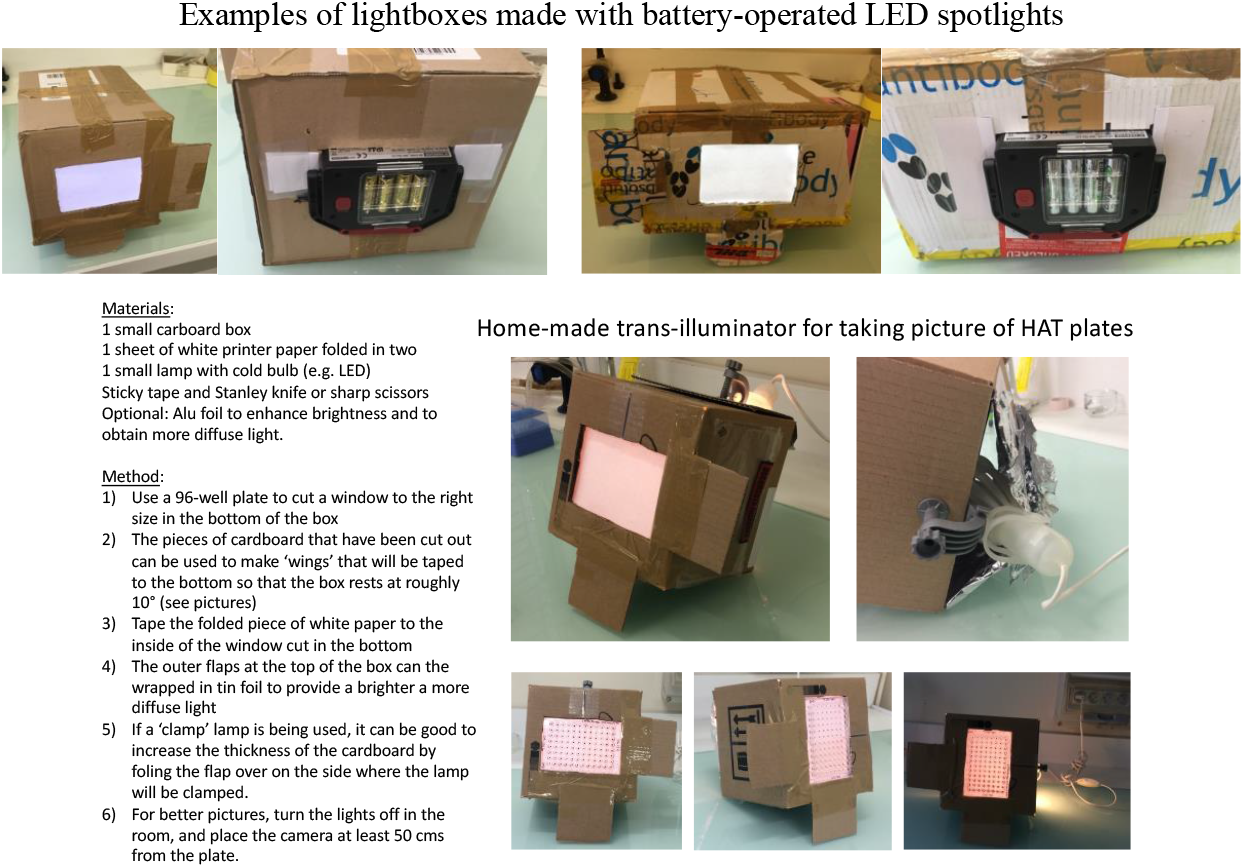

The refereeing process of this manuscript was handled by Review Commons (https://www.reviewcommons.org). The helpful comments and suggestions from three referees have led us to modify our manuscript, and this pdf contains de modified version.

The comments from the three referees, and our point by point replies, are provided below.

------------------------------------------------------------------------------

**Reviewer #1** (Evidence, reproducibility and clarity (Required)):

In the publication HAT-field: a very cheap, robust and quantitative point-of-care serological test for Covid-19 by Joly and Ribes the authors describe an adaption and an improved protocol to their previously published haemagglutination based test to detect antibodies to SARS-CoV-2 in patient blood (Towsend et al., 2021). In detail, they analyzed the effect of several adaptions including buffer optimization, plate coating, usage of patient whole blood instead of washed RBCs and plasma. Additionally they tested different temperatures and stability of the reagents, namely the nanobody-RBD construct IH4-RBD. For validation they compared their optimized HAT-field assay with Jurkat-S&R as a FACS-based assay.

<u>Major comments:</u>

Introduction: This section is rather short and could benefit from a broader overview of currently established methods and assays to detect appropriate immune responses against SARS-CoV-2. The author are advised to summarize the current literature in the field more comprehensively and not only focus on their own work.

> *<u>Response</u>*: *Hundreds of different tests to monitor immune responses against SARS-CoV-2 have been described to date, and the literature on these various tests is vast, with new articles coming out almost on a daily basis. We would not feel either that the introduction of our rather technical paper would benefit from being lengthened by such a review of the current literature, or even competent to carry out such a summary. Following the referee’s suggestion, we have, however, introduced a new sentence and given three references providing relatively recent overviews on the subject of immune-monitoring*.

Cross-reactivity with IH4-RBD.

In Figure 6, the authors highlight the samples in red and orange that showed cross-reactivity with IH4-RBD. In their discussion, however, the authors state that only 2 of 60 (3%) were cross-reactive. In making this statement, they ignore the proportion of cross-reactive samples that were also positive in the Jurkat S&R assay. Therefore, the authors should acknowledge in the discussion that the actual number of cross-reactive samples was higher.

> *<u>Response</u>*: *The statement in the discussion about 2 cross reactive samples out of 60 concerns the results obtained after an incubation of one hour under normal gravity, and not the two red dots in each of the three graphs of figure 6, which correspond to the two negative samples which gave false-positive results in HAT plasma titrations after spinning (Figure 6C), for which we correctly state in the discussion that 12 samples showed cross-reactivity on IH4 alone. The data presented in Figure 6B corresponds to HAT-field after spinning, for which we correctly state in the discussion that 5 out of 60 showed cross-reactivity (4 orange dots + 1 red dot, the second red dot having a score of 0, in accordance with the fact that this sample showed no cross reaction on IH4 alone in HAT-field after spinning)*.
>
> *To try to prevent this possible confusion, we have now clarified what data we are referring to at the start of that paragraph in the discussion*.

Quantitative Assay.

Since the HAT assay does not allow determination of the absolute number of antibodies reactive to SARS-CoV-2 in the blood samples, the authors should refrain from claiming that the HAT-field is a quantitative assay.

> *<u>Response</u>*: *Since immune sera are inherently polyclonal, they contain a multitude of different types of antibodies of different affinities and avidities, and we are not aware of any technique that allows to determine the “absolute number” of antibodies directed against a given antigen in such samples. For many serological tests, including ELISA and the initial protocol of HAT, serum or plasma titrations are used as a means to obtain what is widely considered as a quantitative evaluation of the amounts of antibodies in blood samples. Even FACS-based assays such as the Jurkat-S&R-flow test we have used, are commonly considered as quantitative but those only provide relative results and not absolute numbers. We perceive that the close correlations we find between the results of the HAT-field protocol and those of the Jurkat-S&R-flow test as well as with serum titrations using the standard HAT protocol warrants considering the results of HAT-field as being as quantitative as those obtained with all those other tests*.

Morphological read out

For field application, the morphological description of the observed deposits (“teardrop” vs. “button”) could be problematic and might lead to bias depending on the user. Thus, the authors should provide a clearer description for phenotype classification.

> *<u>Response</u>*: *We have now introduced a specific paragraph detailing how to score HAT assays in the Methods section, as well as a new figure providing a graphic description of positive, partial and negative RBCs deposits*.

<u>Minor comments:</u>

Title: the authors should remove “very”

> *<u>Response</u>*: *We have now removed the word ‘very’ from the title, and thank the referee for this helpful suggestion*.

By the way: What are the costs of IH4-RBD for a 96 well plate? Who will produce this reagent? Is the sequence of the IH4 fully disclosed?

> *<u>Response</u>*: *As specified in our original paper (see Townsend et al. 2021), the plasmid coding for the IH4-RBD is available upon request from Alain Townsend (Oxford, UK). Furthermore, his laboratory funded the production of 1 gram of the IH4-RBD reagent by a commercial company, and professor Townsend has been graciously sending aliquots of 1 mg of this reagent, which suffice for several thousand tests, to all the laboratories that have requested it from him*.
>
> *In its initial format, HAT only required 100 ng of IH4-RBD per well, corresponding to a cost of 0.0027 £ per well. For the HAT-field protocol, 5 times more reagent is needed, thus bringing the cost of the reagent to 1.5 cts per test, to which one would have to add a similar cost for the IH4-reagent alone. This would thus bring the cost of the two reagents to approximately 3 cts, which is still lower than the price of any of the cheap disposable plasticware necessary for the test (lancet, pipet, plastic tube and portion of a plate).*
>
> *The sequence of the IH4 nanobody is indeed fully disclosed (see figure 1 of Townsend et al. 2021), and has actually been protected by a patent (US9879090B2). Whilst IH4 can be used freely for research purposes, licensing rights would have to be taken into consideration by any health authority wishing to use the technique broadly, or for any commercial distribution*.

The usage of the CR3022 as positive control for neutralizing antibodies should be reconsidered since this antibody does not confer viral neutralization. Other well describe antibodies blocking the ACE2:RBD interface might be better suited.

> *<u>Response</u>*: *CR3022 was the one that we had at our disposal, but other mAbs can certainly be used instead of as positive controls, and this is actually indicated in the detailed HAT-field protocol provided. Since the use of a positive control is only to ensure that the IH4-RBD has not been degraded and works as well as expected, and that any negative samples are not due to a very rare glycophorin mutation that could prevent IH4 from binding to it at the surface of RBCs, we are not sure why using a mAb with neutralizing activity would necessarily be better than the CR3022 mAb*.

Figure 2: Please state the concentration of IH4-RBD used. As stated in the figure legends for Figure 2 B, the authors should show the result all 4 replicates (incl. SD)

> *<u>Response</u>*: *The concentration of IH4-RBD was 1* μ*g/ml, i*.*e. the normal concentration for standard HAT tests. This was already indicated in the Methods section, but has now been added to the legend of Figure 2*.
>
> *Whilst 4 experiments were indeed carried out, which all gave similar results, i*.*e. showed that using PBS-N3 or PBN did not hinder HAT performance, but could instead result in a slight increase in HAT sensitivity, those various experiments were not all exact replicates of the experiment shown on figure 2. Furthermore, performing of those various experiments was spread over a period of over a year, using different reagents, thus precluding numerical comparisons between the various results. We have clarified this issue by rewording the final statement to “Comparable results were obtained in four similar experiments*.*”*

Figure 3: Although the authors showed stability of IH4-RBD at 2 μg/ml they do not provide data for the stabilities at higher dilutions. As the authors suggest to predistribute the IH4-RBD in plates they should at least discuss this issue.

> *We thank the referee for raising this valid point, which has now been discussed in the paragraph entitled “Practical considerations for performing HAT assays” in the Methods section: “One aspect that will have to be considered for the design and use of such individual strips of wells will be to ensure that, upon storage, the various dilutions of IH4-RBD are as stable in such strips as the working stocks of IH4-RBD (2* μ*g/ml) tested in Figure 3*.*”*

Figure 6/Supplementary Figure 1 and 3

The presentation of the data is not accurate, as many of the points (samples) are obviously identically positioned in the graph. The authors should choose a different representation of their data. E.g. they could adjust the size of the points to the number of overlapping samples.

> *<u>Response</u>*: *We thank the referee for raising this issue, which was also pointed to by referee #2. This apparent inaccuracy is due to the fact that, on these plots, the scales for both x and Y axes used discrete values, which indeed results in multiple points overlapping on top of one another. This was resolved by adding numbers next to the positions where several dots overlapped*

Wording / text length

In the current manuscript the text is very long. Thus, the authors should shorten it to report the essential findings more appropriately. Additionally they should check for correct English wording.

> *<u>Response</u>*: *We thank the referee for this remark, which helped us realize that the excessive length of the manuscript was mostly due to an extensive discussion of highly technical and practical points. The corresponding paragraphs were indeed out of place in the general discussion, and have not been deleted but have been moved to the Methods section since we feel that they contain very important information for people who would actually start to performing HAT assays*.

<u>Reviewer #1 (Significance (Required)):</u>

In summary, the authors describe the HAT-field test as a simple PoC test for the detection of SARS-CoV-2 antibodies in patients. Because of its ease of use and robustness, the test appears to be particularly well suited for use in countries with underdeveloped health care or limited testing facilities, as also reported previously. The value of this manuscript lies mainly in the detailed description of the protocol and its validation. In this context, the adaptations described are certainly useful and helpful from a practical point of view, but do not provide significant new scientific insights. In light of these considerations, we recommend that this work be submitted to an appropriate journal specializing in the publication of such methods

<u>Expertise</u>

The reviewers have established and published different serological assays to monitor immune responses against SARS-CoV-2

**Reviewer #2** (Evidence, reproducibility and clarity (Required)):

In this paper, the authors developed a feasible protocol for an affordable point-of-care serological test for SARS-CoV-2. This method was adapted from the HAT plasma titration test that the authors previously published. Specifically, the test utilizes a 96-well plate pre-coated with the RBD of SARS-CoV-2 spike glycoprotein fusing to a red blood cell targeting nanobody (IH4). By adding microliters amount of the blood or plasma samples to the plate, it allows the detection of antibodies against RBD by measuring the level of hemagglutination. In the current upgraded protocol (so called HAT-field), the authors made major modifications including optimizations of buffer and experimental protocol and the use of pre-titrated IH4-RBD on the plate, which collectively helped to lower the sample consumptions, improved the stability and the sensitivity of detection, and made the test more user-friendly under non-clinical settings.

<u>Major comments:</u>

My major concerns are related to the robustness and quantitative capability of this approach. Specifically:

It seems that multiple variables may impact the results. These include volume of droplets, the presence/absence of serum IH4 or BSA cross-reactive antibodies, and the amount (%) of red blood cells which may vary substantially among samples. Could you find a way to normalize the results (e.g., the discrepancy shown in Figure 6) instead of only leaving them as false-positives or false-negative?

> *<u>Response</u>*: *Regarding the volume of the droplets, in other words, the amount of blood collected and used in an assay, two sentences in the manuscript underline the fact that this is not a critical variable:*
>
> *In the Results section “the precise volume of blood collected is not critical; it may vary by as much as 30% with no detectable influence on the results*.*”*
>
> *In the discussion: “On this subject, we have found that increasing the amount of whole blood per well (in other words using blood that is less dilute) has very little influence over the HAT-field results, and, if anything, adding more blood can sometimes reduce the sensitivity, albeit never by more than 1 dilution*.*”*
>
> *Consequently the % of RBCs in samples seem unlikely to influence the HAT-field scores significantly. This is supported by the fact that, although men tend to have higher hematocrits than women, we have not noticed any detectable difference between men and women in the correlation of the HAT-field scores with those of the Jurkat-S&R-flow test*.
>
> *We are not sure that we fully understand what discrepancy shown in Figure 6 the referee is pointing to, but if it is about the increase in the number of samples found to be cross reacting on IH4 alone when the sensitivity increases, in the discussion, we propose to perform tests using titrations of the IH4 nanobody alone simultaneously to using the IH4-RBD reagent, so as to minimize the number of samples that would be identified as false positives if only one concentration of IH4 alone was used as negative control. Comparing the titers obtained with IH4-RBD and IH4 alone will then provide some level of normalization for the samples cross reacting on IH4. As for the hypothetical presence of antibodies cross reacting on BSA alluded to by the referee, since such antibodies would not bind to RBCs, we do not think they would affect the HAT results*.

Second, the score of the HAT-field ranges from 0 - 8. However, based on the current manuscript, it is not clear how the scoring and scaling works. How is the noise (non-specific antibody signal) defined here?

> *<u>Response</u>*: *We have now introduced a specific paragraph and a new figure detailing how to score HAT assays in the Methods section*.

In addition, it is unclear how to translate the HAT-field score into a meaningful measure of protection by serum antibodies.

> *<u>Response</u>*: *Documenting the correlation between HAT-field scores and levels of protection against SARS-CoV-2 infections and/or Covid-19 severity would indeed be extremely interesting. This would, however, require setting up a large scale clinical trial carried out over several months. This type of work could only be carried out by a large consortium including clinicians or even preferably a national health agency. This was, however, far beyond the reach of this initial project, which was based on the work of a single person on a shoestring budget*.

Can you provide more evidence to demonstrate that the test is quantitative? For example, performing additional orthogonal experiments to better validate the scoring and generate a correlation function?

> *<u>Response</u>*: *Inasmuch as it would have been very interesting to perform additional serological tests from commercial sources on the samples of our cohort, such tests are all very expensive (e*.*g. ca. 500 € for one ELISA plate). This was in fact the main reason for developing the Jurkat-S&R-flow test in the first place, since it is much cheaper, more modular, and at least as sensitive as ELISA (see Maurel Ribes et al. 2021). The funds for this whole project came from a single 15 k€ grant obtained from the ANR, and we simply did not have access to the funds, or to the human resources to carry out such experiments based on commercial serological tests*.

<u>Minor comments:</u>

Figure 6: are all results included? To me, it does not seem that all 60 samples data were included in the plot.

> *<u>Response</u>*: *We thank the referee for raising this issue, which was also pointed to by referee #1. This apparent inaccuracy is due to the fact that the scales for both x and Y axes used discrete values, which results in multiple points overlapping on top of one another. This was resolved by adding numbers next to the positions where several dots overlapped*.

There are several redundant statements in the discussion and results section. Please make the text more concise.

> *<u>Response</u>*: *The discussion has now been shortened considerably, mostly by moving the paragraphs pertaining to technical considerations to the Methods section*.

<u>Reviewer #2 (Significance (Required)):</u>

The current paper is built upon the improvement of previous published work. In addition, there are similar approaches that have been published. It was unclear if the current method is superior to other works.

> *<u>Response</u>*: *Whilst we have made no statement regarding whether the method we describe is superior to other methods, we are pretty confident that very few alternatives will be as frugal and simple as the HAT-field protocol described here. As alluded to in the final paragraph of the discussion, two recent reports have described that HAT could be performed on cards rather than in V-shaped wells, with semi-quantitative results being obtained in minutes. If such card-based approaches turn out to provide sensitivity and reliability comparable to those of the HAT-field protocol, they will certainly represent very interesting alternatives. As stated in our manuscript, we would be very interested if the comparative evaluation of the two approaches could be carried out by one or several independent third party*.

My research involves the development of antiviral antibody therapeutics. This method may be used as a point-of-care tool for the measurement of serologic response to RBD in less developed countries. However, due to the high vaccination rate and large infected populations, the overall needs for such detection drastically decrease. The significance of the work and utilities of the test may expand with more experiments related to the variants.

**Reviewer #3** (Evidence, reproducibility and clarity (Required)):

This paper describes a low-cost robust and quantitative serological test based on haemgglutination, which could be used in resource limited settings for evaluating population-based and vaccine induced immunity. Neutralising antibodies to the receptor binding domain (RBD) on the SARS CoV-2 spike protein are an immunological correlate of protection. The HAT has a single reagent the RBD domain of SARS CoV-2 linked to a monomeric anti-erythrocyte single domain nanobody. When human polyclonal serum antibodies bind to the RBD they cross-link and agglutinate human red blood cells, resulting in haemagglutination which can be read visually.

This paper thoroughly evaluate the stability of the HAT reagents used to measure human and monoclonal antibodies examining the robustness of the HAT reagent. It provides a comprehensive protocol for conducting field based HAT with limited reagents. The test can evaluate is subjects have been infected using a simple finger prick to detect RBD specific antibodies. The field HAT can also be used to define people that can be susceptible to reinfection or in need of vaccination, With the use of RBDs from the variants of concern the test can be rapidly adapted to evaluate antibodies as new variants arise to evaluate surrogate correlates of protection to allow timely evaluation of vaccine effectiveness and predict the need for vaccine booster doses.

The data are very comprehensively presented with good figures demonstrating the most appropriate buffer to store the IH4-RBD reagent and the robustness of the HAT over time at different temperatures.

No additional experiments are needed and suitable numbers of replicates are included. All data, methods and reagents are comprehensively described.

<u>Minor comments:</u>

The paper is well written but rather long in places and may have benefited from being more succinct.

> *<u>Response</u>*: *The excessive length of the manuscript was mostly due to an extensive discussion of highly technical and practical points. The discussion has now been shortened considerably, mostly by moving the paragraphs pertaining to technical considerations to the Methods section*.

Panels in figures could be labelled as A, B, C etc to help in identifying the correct panel..

> *<u>Response</u>*: *We thank the referee for this helpful suggestion, which we have followed*.

I would avoid the use of experiment and project and refer to next we confirmed… or in this paper or our results show

Please make sure all abbreviation are defined upon first use.

Perhaps include early in the paper that most of the work was conducted with the Wuhan RBD

> *<u>Response</u>*: *We thank the referee for these helpful suggestions, which we have followed to the best of our abilities. The abstract now contains a mention of the fact the work on optimizing the protocol was carried out with the IH4-RBD carrying the Wuhan version*.

Figure 2: I would suggest placing either a solid line between the two halves of the plates to make it easier for the reader to differentiate between the two antibodies. It also would have been easier to read if the bottom PBS, PBS-N3 and PBN were at 45 degree angle. In B include the serum name (e.g. serum 197).

> *<u>Response</u>*: *We thank the referee for these helpful suggestions, which we have followed*.

Legend to figure 4: please include the serum numbers after covid-19 patients. Perhaps include arrows to demonstrate the dilutions of serum and IH4-RBD in the figure.

Page 6 it might be easiest to use the same times as in figure 6 and use for example more than one year in the discussion

> *<u>Response</u>*: *We thank the referee for these helpful suggestions, which we have all followed*.

Legend figure 6 perhaps replace dots with circles page 10 include the R values from figure 6 in the description of results.

> *<u>Response</u>*: *We are grateful to the referee for these helpful suggestions, but have not followed them since we do not feel that these changes would be real improvements*.

Page 12 of note perhaps this can be moved to the methods ?

> *<u>Response</u>*: *This, and several other paragraphs of the Discussion, have now been moved to the Methods section*.

Supplementary figure 2 A can be seen, is something missing here?

> *<u>Response</u>*: *An s was indeed missing* : *“A can be seen” corrected to “As can be seen “*

<u>Reviewer #3 (Significance (Required))</u>:

This paper describes a simple rapid field test for evaluating antibodies to the receptor binding domain of the spike of SARS CoV-2 using the Wuhan and delta variant. Whilst high income countries can provide booster doses and extensive testing (either lateral flow or RT-PCR based) and contact racing to control the waves of the pandemic, low income countries have had limited access to Covid vaccine and the extent of previous waves of the pandemic in the populations are unknown.

This paper describes a robust and simple test for investigating human antibodies to SARS-CoV-2 which could be performed in resource limited settings providing a very useful tool for monitoring infection in the community and potentially for prioritising this scarce COVID-19 vaccines available.

This study builds upon the work conducted on the HAT and has extensively studied and optimised the test so that it could be used globally. This paper provides a comprehensive protocol and has simplified the test to ensure it could be used in LMICs.

This paper would be of great interest to a wide scientific audience who are interested in a rapid low-cost test to evaluate population based and vaccine induced immunity.

<u>Reviewer #3 Expertise</u>: serological assays for use in virology and vaccinology. Suitable competence to review the whole paper

